# Estimating Impact of SARS-CoV-2 Infection on Health-Related Quality of Life among Persons Aged 8 years and Older, August 2020-July 2022

**DOI:** 10.64898/2025.12.10.25341971

**Authors:** Sheroi Johnson, Huong Q. Nguyen, Melissa S. Stockwell, Christina A. Porucznik, Ruth A. Karron, Fatimah S. Dawood, Lisa A. Prosser, Alisha P. Sarakki, Joseph B. Stanford, Marissa Hetrich, Elizabeth Schappell, Maria Deloria Knoll, Vic Veguilla, Alexandra Mellis, Jazmin Duque, Zuha Jeddy, Jamison Pike, Melissa A. Rolfes

## Abstract

**Background:** Severe COVID-19 results in substantial economic burden and impacts quality of life. Assessing how non-hospitalized COVID-19 impacts health utilities during acute infection and long term is important to estimate the full economic impact of SARS-CoV-2 infection.

**Methods:** We analyzed EQ-5D-3L survey data from SARS-CoV-2 infected adults (aged ≥16 years) and children (aged 8-15 years) from three community and household cohorts in the United States (2020-2022). EQ-5D-3L scores were analyzed at three time points after symptom onset or first positive SARS-CoV-2 test result and converted to health utilities on a scale of 0-1 (1=perfect health). Among adults, regression models were used to compare differences in health utility by demographic/clinical characteristics.

**Results:** Among 538 SARS-CoV-2 non-hospitalized asymptomatic/symptomatic infections from 575 adults with EQ-5D-3L surveys, mean utilities were near 1 throughout the observation period. During 0-14 days, vaccinated participants had higher health utilities (Beta:0.57, 95% CI:0.07,1.07). Seeking medical care and having gastrointestinal symptoms (vs. none), were associated with lower health utilities (Beta, 95% CI:-0.96, −1.60, −0.31; and −0.76, −1.30, −0.21 respectively). During 15-30 days, unemployment was associated with lower health utility (Beta:-0.64, 95% CI:-1.15,-0.14). During 31-90 days, underlying conditions were associated with lower health utilities (Beta:-0.32, 95% CI:-0.54, −0.09). Results for children were similar to adults.

**Conclusion:** Non-hospitalized COVID-19 may have minimal overall impact on quality of life; however, health utilities differed by vaccination status, presence of gastrointestinal symptoms, employment status, and presence of underlying conditions. Vaccination may play an important role in minimizing illness impact from SARS-CoV-2 infection.

**Key points:**

- Severe COVID-19 illness causes substantial economic burden and impacts on quality of life; however, the incidence of non-hospitalized COVID-19 is far greater. Assessing how non-hospitalized COVID-19 impacts health during acute infection and long term is important to understand the full impact of SARS-CoV-2 infection.
- This study utilizes the EQ-5D-3L, a standardized generic preference-based instrument used in population health studies, to estimate health utilities at multiple time points following SARS-CoV-2 infection. The study also examines demographic/medical characteristics that are associated with health utility over time.
- While mean health utilities were high for all infection periods regardless of age, health utility was lower at varying time points post-infection among adults who sought medical care, reported gastrointestinal symptoms, were unemployed, and with underlying conditions. Vaccinated adults (vs. unvaccinated) had higher health utility and were less likely to report reduced health. Findings can be used as inputs for economic evaluation and assessing impact of interventions for non-hospitalized SARS-CoV-2 illness, such as vaccination.

## Introduction

Since the first detected infection in the United States (U.S.) in January 2020 [1] the COVID-19 pandemic, caused by the SARS-CoV-2 virus, has led to over 1 million hospitalizations and deaths in the country.[2, 3] Severe COVID-19 illnesses represent substantial economic burden and impacts on health;[4, 5] however, the incidence of non-hospitalized COVID-19 illnesses far exceeds that of severe illnesses.[6, 7] Nonetheless, non-hospitalized COVID-19 can still have a negative impact on a person’s quality of life and because they are far more common than hospitalized illnesses, the combined negative impact may be large and important to assess, especially among those with long COVID, where symptoms may persist for at least 3 months after infection.[8, 9] Estimating how SARS-CoV-2 infection impacts a person’s quality of life during acute infection and in the long term, is important to fully characterize the burden of COVID-19 and inform assessments of the value of prevention and mitigation interventions.

Previous studies of impact of COVID-19 on health-related quality of life have used the EuroQol five-dimension questionnaire (EQ-5D), a standardized generic preference-based instrument used to measure self-rated health,[7, 10] and have reported declines in health-related quality of life among non-hospitalized persons with mild illness[11–13] as well as hospitalized patients with severe illness.[14–16] We build upon this evidence to estimate health-related quality of life among individuals with asymptomatic to symptomatic, non-hospitalized SARS-CoV-2 infection identified from community and household cohorts with systematic testing for SARS-CoV-2. Using these cohorts, we captured participant health prospectively at enrollment and at defined periods after SARS-CoV-2 infection to calculate health utilities, assess changes from enrollment in self-reported health, and examine differences by demographic and medical characteristics. This is unique to our study design as previous studies estimating health-related quality of life associated with COVID-19 infections were retrospectively conducted. Another unique and important attribute of this study is use of a serial cross-sectional approach to capture health at multiple time points after infection rather than a single time point.

## Methods

### Source population

This analysis included data from three prospective, longitudinal COVID-19 cohorts within the U.S. followed between 2020 and 2022: 1) Coronavirus Household Evaluation and Respiratory Testing (C-HEaRT), 2) Prospective Assessment of COVID-19 in a Community (PACC), and 3) SARS-CoV-2 Epidemiology And Response in Children (SEARCh). The C-HEaRT cohort (N=1485 people) recruited and followed households with at least one child aged <18 years from Salt Lake City, Utah from August 2020 to August 2021, and New York City, New York from September 2020 to August 2021.[17] The PACC cohort (N=1520 people) randomly sampled and enrolled community members near Marshfield, Wisconsin and followed participants from November 2020 to July 2022.[18] The SEARCh cohort (N=759 people) recruited households with at least one child aged <5 years in Maryland and included follow-up from November 2020 to October 2021.[17] In each cohort, all participants completed an enrollment survey collecting demographic/medical information followed by weekly symptom assessments collecting information on presence of fever/chills, cough, loss/change in taste/smell, sore throat, muscle/body aches, shortness of breath/difficulty breathing, diarrhea, fatigue, headache, nasal congestion, and vomiting and weekly self-collected respiratory specimens regardless of symptom status. All participants were asked to assess their current health at enrollment and end of follow-up by completing a general health survey consisting of two parts: 1) visual analog scale (VAS; ranking their health on a scale from 0 [worst health imaginable] to 100 [best health imaginable]) and 2) EQ-5D-3L.[10, 19] Responses from EQ-5D-3L were used to compute health utilities, which were assessed for correlation with VAS scores. Participants in the PACC cohort completed the EQ-5D-3L every 6 months as well as the end of the follow-up period. Participants with a respiratory specimen positive for SARS-CoV-2 during the study period were asked to complete the EQ-5D-3L and VAS after the earliest of either symptom resolution (defined as ≥3 days without COVID-19-like illness symptoms) or 4 (PACC cohort) or 5 weeks (C-HeaRT and SEARCh cohorts) for every positive specimen. For C-HeaRT and SEARCh cohorts, EQ-5D-3L/VAS illness follow-up surveys inquired about health during illness. For the PACC cohort, EQ-5D-3L/VAS illness follow-up surveys were administered to collect information about current health. Participants were given the option to complete all data collection surveys online or via telephone.

This study was reviewed and approved by the Institutional Review Boards at the Marshfield Clinic Research Institute (PACC), the University of Utah and Columbia University (C-HEaRT), and Johns Hopkins University (SEARCh). The Centers for Disease Control and Prevention Institutional Review Board reviewed these activities and relied on the approvals of the other participating institutions (see 45 C.F.R. part 46; 21 C.F.R. part 56).

### Description of EQ-5D-3L survey and Health Utility

All EQ-5D-3L responses were converted to health utilities at key time points for further analysis: at enrollment, every 6 months (in the PACC cohort), end of follow-up, and after SARS-CoV-2 infection. Health utilities are values that represent the strength of an individual’s preference for specific health-related outcomes with values ranging from 1 (equal to perfect health) to 0 (equal to being dead); values can also be negative (indicating worse than being dead).[10] The EQ-5D-3L asks individuals to rate their current health across five domains (mobility, self-care, usual activities, pain/discomfort, and anxiety/depression) using a scale from 1 (no problems) to 3 (extreme problems). The completed 5-domain response to the EQ-5D-3L is known as a health state.[19] Value sets (or preferences) for each health state have been solicited from the general population in various countries.[20–22] Health utilities for adults (aged ≥16 years) in this study’s cohorts were calculated using previously published U.S. value sets, elicited using the time trade-off valuation method and then mapped to EQ-5D-3L profiles.[20] As no U.S. value sets existed for children aged 8-15 years, value sets from Spanish children using a time trade-off valuation were used.[23] No value sets existed for children aged <8 years old, thus cohort participants aged 4-7 years were excluded from the analysis.

### Demographic/medical characteristics

Participants self-reported demographic characteristics at enrollment, including age, sex, race/ethnicity, highest educational level attained, and current employment: employed (full/part-time) and unemployed. Participants also self-reported the presence of select underlying conditions at enrollment, including obesity (derived from self-reported height and weight), mental disorders, lung disease, liver disease, kidney disease, immunocompromised disease, hypertension, heart disease, diabetes, chronic obstructive pulmonary disease (COPD), and cancer. SARS-CoV-2 vaccination status was assessed throughout each cohort’s study period. For C-HEaRT and SEARCh cohorts, vaccination information was self-reported at enrollment and end of follow-up and verified with COVID-19 vaccination cards or data from local immunization information systems or registries. For the PACC cohort, vaccination information was acquired through electronic health records and the Wisconsin Immunization Registry. Individuals’ vaccination status categories were classified by vaccination status at time of their SARS-CoV-2 infection: unvaccinated (no doses of SARS-CoV-2 vaccine), partially vaccinated (1 dose of a 2-dose SARS-CoV-2 vaccine), or fully vaccinated (2 doses of 2 dose SARS-CoV-2 vaccine or 1 dose of 1 dose SARS-CoV-2 vaccine).

For analysis, symptoms were grouped into eight syndromes: influenza-like illness (ILI; fever/chills and cough or sore throat), COVID-like illness (CLI; fever/chills, cough, or shortness of breath/difficulty breathing), upper respiratory symptoms (nasal congestion or sore throat), lower respiratory symptoms (cough or shortness of breath/difficulty breathing), neurological symptoms (headache or loss/change in taste/smell), gastrointestinal symptoms (vomiting or diarrhea), constitutional symptoms (fever/chills, muscle/body aches, or fatigue), and moderate COVID-19 symptoms (fever/chills or shortness of breath/difficulty breathing).[24–26] Individuals could experience one or more syndromes during any illness episode. Surveys after SARS-CoV-2 infection captured whether participants sought medical care for their illness in the 30 days after onset of symptoms or their positive test.

### Statistical analysis

Data analyses were performed using R (version 4.1.0)[27] using the package “eq5d”.[28]

#### Analytic population

The analytic population included non-hospitalized participants with a PCR-positive SARS-CoV-2 test result during the study follow-up period, and who had at least one completed EQ-5D-3L survey in the 90 days after SARS-CoV-2 infection. Three post-SARS-CoV-2 infection periods were defined for analysis: 0-14 days after onset/positive test (called “early period”), 15-30 days (“middle period”), and 31-90 days (“late period”). Participants aged ³16 years were categorized as adults and those aged 8-15 years were categorized as children. While in many legal and public health frameworks, 16- and 17-year-old individuals are considered as children, the EQ-5D-3L adult version is recommended for those aged ≥16 years[19], with health state preferences of older adolescents being more comparable to adults than younger children.[29]

#### Health utility

For each post-SARS-CoV-2 infection period, the distributions of health utilities were examined overall and by demographic or medical characteristics. Correlation between VAS scores and calculated healthy utilities at the same time point was also examined. Among adults, beta regression models were used to assess whether there were differences in health utility by demographic or medical characteristics. A beta distribution was chosen because of its similarities to the distribution of health utilities in which the values range between 0 and 1 (not inclusive) and highly skewed distributions are allowed.[30] For beta regression, health utility values of 1 were transformed by the formula: Y* = [y(N-1) + 0.5]/N, where Y* is the transformed health utility, y is the original health utility, and N is the sample size,[30] yielding values of 0.9959. There were no health utilities with a value of 0 in this analysis. With the exception of models specifically assessing the association between age and health utility, all models were adjusted for age (as a continuous variable). Models assessing education and employment were restricted to adults aged ≥18 years.

#### Health change

The Paretian Classification of Health Change (PCHC)[10] was used to compare changes in domain-specific responses from a participant over time, comparing each EQ-5D-3L response at enrollment to the response during each infection period. The PCHC categorizes health state changes over time as: *No Change, Improve, Worsen, Mixed Change.*[10] A health state is “improved” if it is better on at least one domain and no worse on any other domain, “worsened” if it is worse in one domain and no better in any other, and “mixed” if it is better in at least one domain, but worse in at least one other.[10] PCHC analyses were restricted to participants with SARS-CoV-2 infection who completed a survey at enrollment and at least one survey ≤90 days after SARS-CoV-2. For each infection period, the earliest survey after symptom onset/positive test was used in the analysis.

Among adults only, we presented PCHC analyses stratified by demographic/medical characteristics that showed significant associations with health utilities. Fisher exact tests (2-sided) were used to compare the proportion of participants in each health state (*P* values <0.05 were considered statistically significant).

## Results

### Study Population

Across the three cohorts, 3764 participants were enrolled (Figure 1). Participants were excluded if they were aged <8 years or missing age (n=861), did not test positive for SARS-CoV-2 during the study period (n=2136), tested positive for SARS-CoV-2 and were hospitalized (n=3), or with no EQ-5D-3L data within 90 days after symptom onset (n=1) or positive test for SARS-CoV-2 (n=188). The final analysis consisted of 575 participants (469 aged ≥16 years and 106 aged 8-15 years) with SARS-CoV-2 infections detected who completed 662 EQ-5D-3L surveys ≤90 days post-onset/positive test (538 among aged ≥16 years and 124 among aged 8-15 years). Infection dates ranged from October 2020 to June 2022. Out of the 575 participants with at least one SARS-CoV-2 infection, 554 (96%) participants were infected once, and 21 (4%) participants were infected twice. In total, 662 EQ-5D-3L surveys were completed: 94 participants (70 aged ≥16 years and 24 aged 8-15 years) completed the EQ-5D-3L during the early infection period, 144 (117 aged ≥16 years and 27 aged 8-15 years) during the middle infection period, and 424 (351 aged ≥16 years and 73 aged 8-15 years) during the late infection period. Eighty-seven (15%) participants completed the EQ-5D-3L twice within the 90 days after infection (8 during early and middle periods; 26 during early and late periods; and 53 during middle and late periods). No participant completed the EQ-5D-3L survey during all three infection periods.

**Figure 1.**
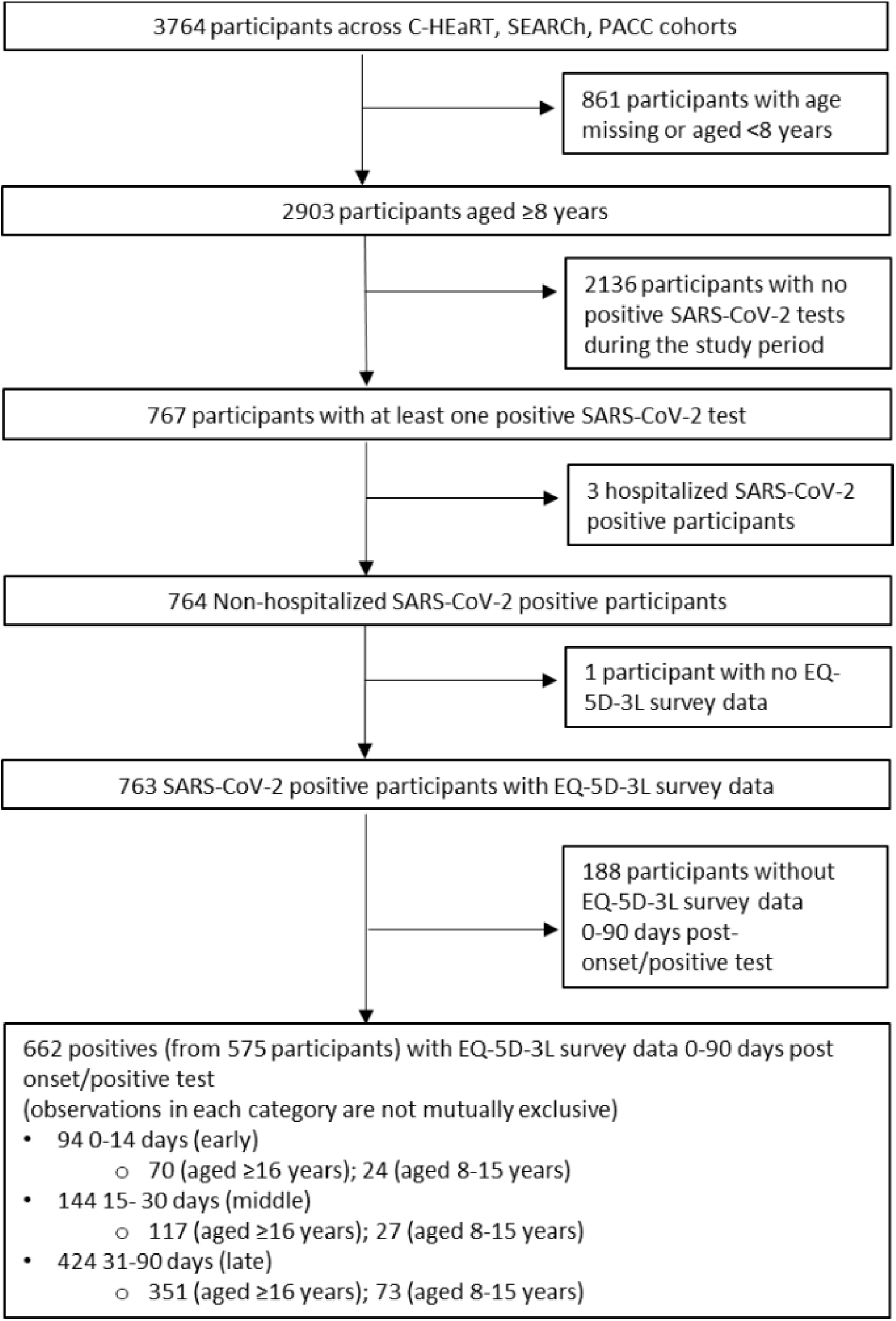
Inclusion and exclusion of participants in analysis of health-related quality of life from three prospective, longitudinal cohorts in the United States.

Among adults in the analysis, most were from the PACC cohort in Wisconsin, female, aged 16-49 years, non-Hispanic White, college graduates, and currently employed (Table 1). Fifty-four percent of the adults with SARS-CoV-2 infection had at least one underlying health condition, most commonly obesity (40%) and hypertension (20%). In the first 30 days following the symptom onset or positive test for SARS-CoV-2, most adults (73%) reported at least one symptom, and 8% sought medical care (Table 1). Fifty-five percent of adults were fully vaccinated prior to their SARS-CoV-2 infection. Demographic and medical characteristics of all participants, including children (aged 8-15 years), with SARS-CoV-2 infection are reported in Supplemental Tables 1 and 2.

**Table 1.**
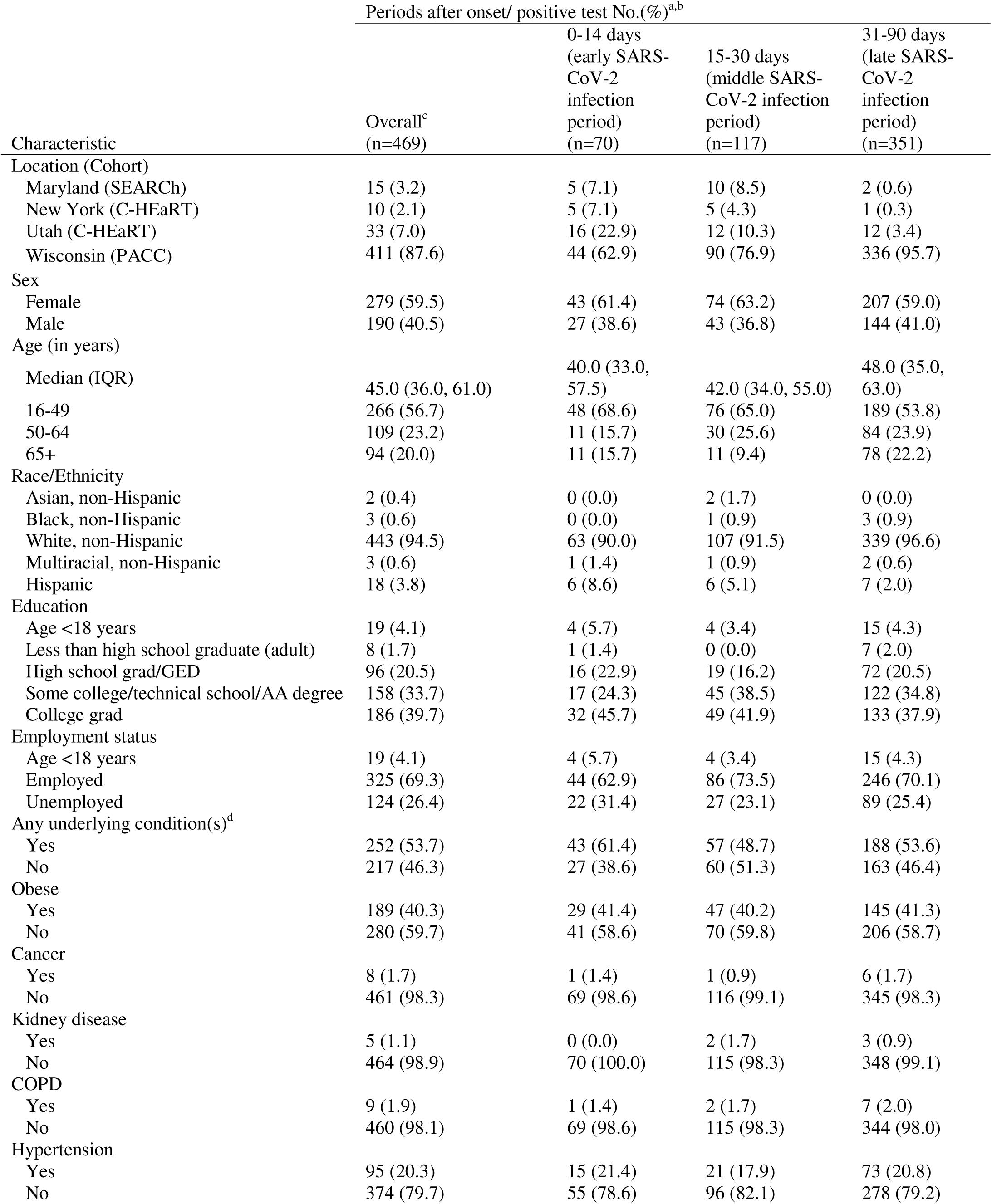
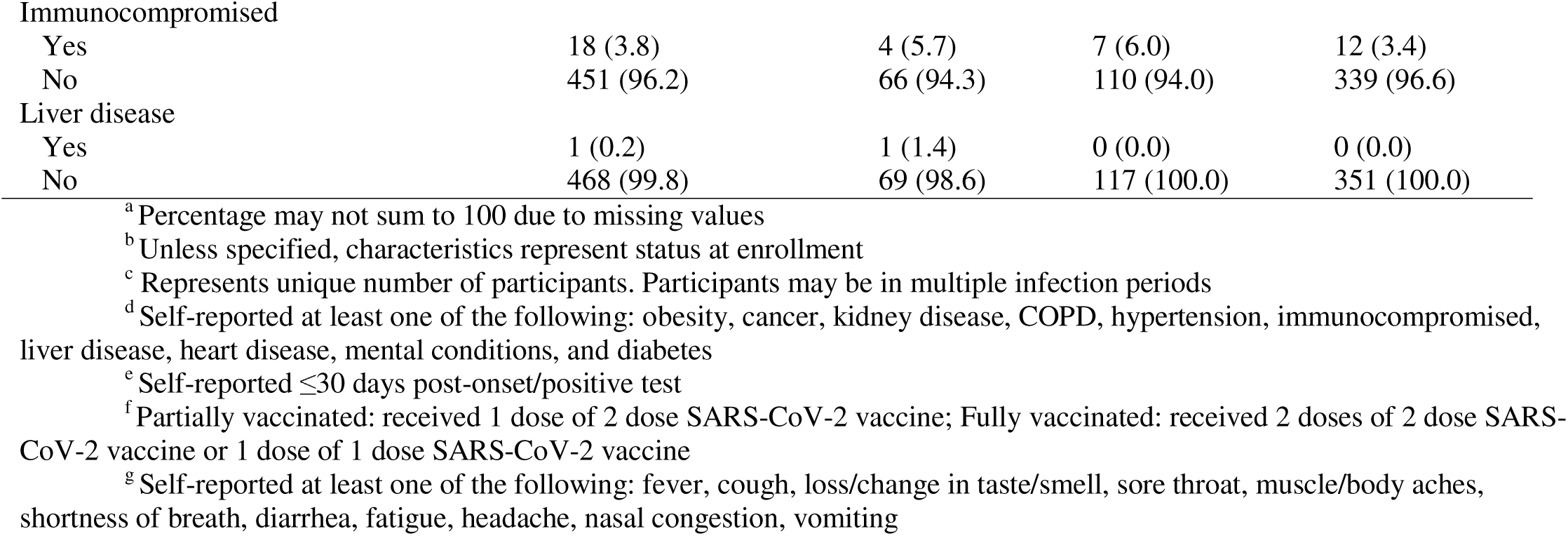
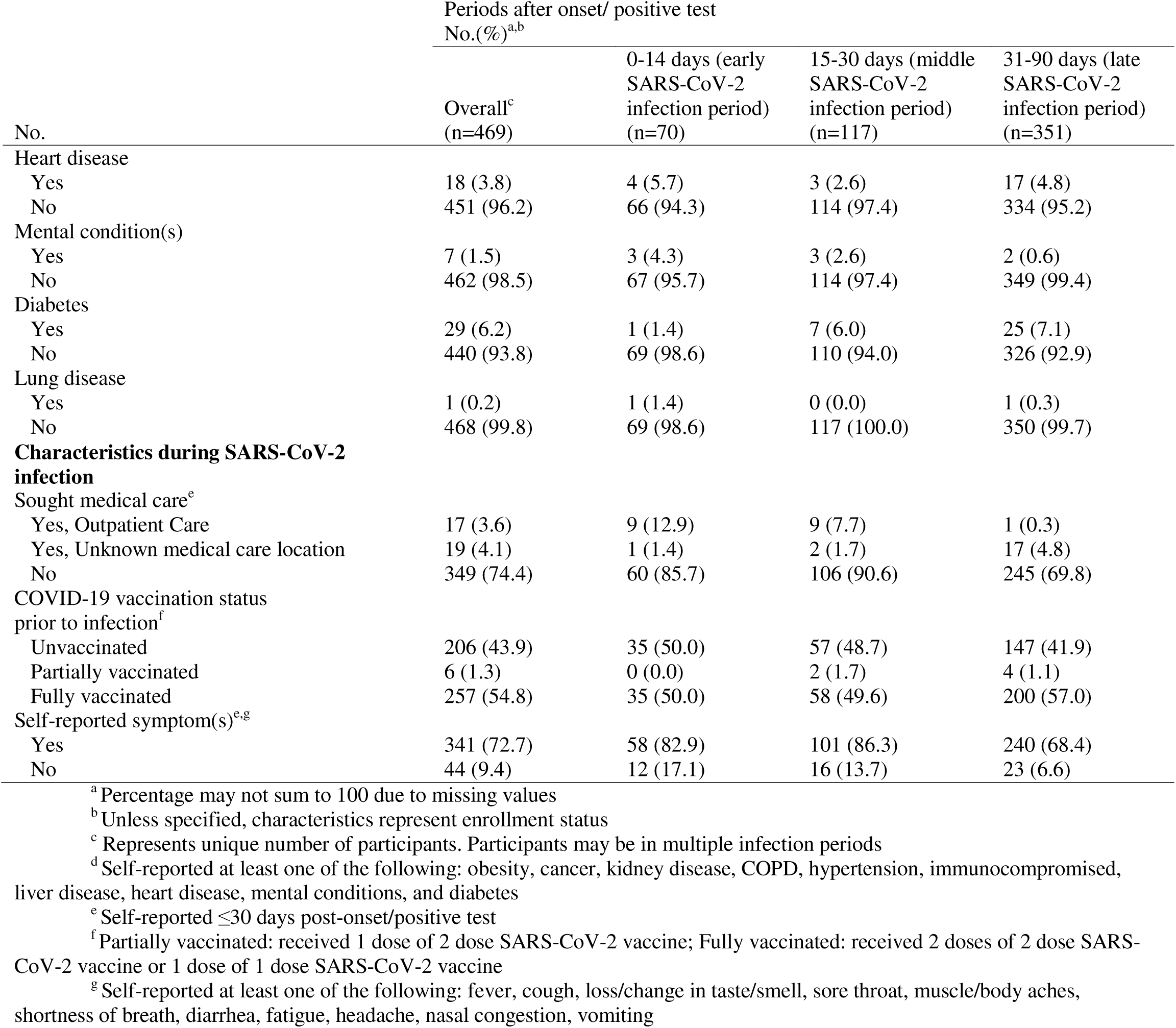
Characteristics of adult participants aged ≥ 16 years with SARS-CoV-2 infection and EQ-5D-3L survey data within 90 days after symptom onset/positive test from three prospective, longitudinal cohorts in the United States from August 2020-July 2022

### Health utilities

Among adults aged ≥16 years, health utilities were skewed towards 1 for all infection periods, with most participants reporting nearly perfect health (Figure 2). However, the mean (SD) and median (IQR) health utility during the early infection period were 0.86 (0.16) and 0.83 (0.80, 1.00) respectively, which were lower, though differences were not statistically significant, than the mean and median during the middle (mean (SD): 0.88 (0.17); median [IQR]: 1.00 [0.82, 1.00]) and late infection periods (mean (SD): 0.91 (0.13); median [IQR]: 1.00 [0.83, 1.00]). Ranges (minimum to maximum) of health utilities were 0.22 to 1.00 (early), 0.17 to 1.00 (middle), and 0.17 to 1.00 (late) (Figure 2 and Supplemental Table 3).

**Figure 2.**
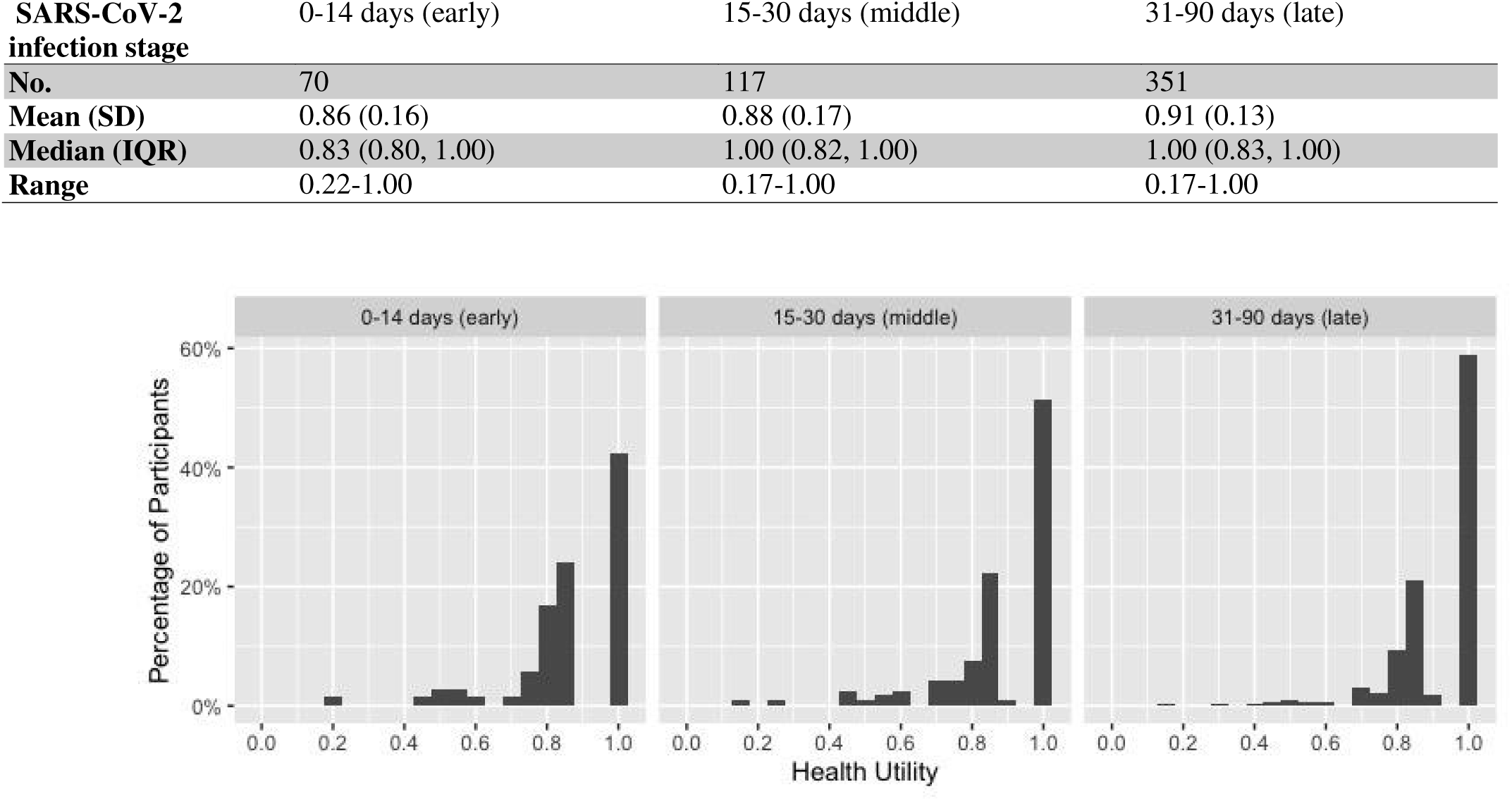
Distribution of health utilities among adults aged ≥16 years with SARS-CoV-2, by infection stage. Health utilities reflect preferences for a specific health-related outcome, with values ranging from 1 (equal to perfect health) to 0 (equal to death). SARS-CoV-2 infection stage was defined as the days after symptom onset/positive test that the EQ-5D-3L survey was completed. SD = standard deviation, IQR = interquartile range.

Among children aged 8-15 years, the mean (SD) and median (IQR) health utility were 0.93 (0.11) and 1.00 (0.89, 1.00) (early); 0.90 (0.20) and 1.00 (0.89, 1.00) (middle); and 0.95 (0.08) and 1.00 (0.89, 1.00) (late) respectively. Ranges (minimum to maximum) of health utilities were 0.61 to 1.00 (early), 0.12 to 1.00 (middle), and 0.71 to 1.00 (late) (Supplemental Table 2). Full descriptions of EQ-5D-3L health states and health utilities in each infection period stratified by age and medical characteristics are presented in Supplemental Tables 3 and 6-11.

Regardless of age, for all infection periods, there was a positive correlation between health utilities and reported VAS scores (Supplemental Figure 1). Correlation coefficients were 0.32 (early), 0.63 (middle), and 0.44 (late) for adults, and 0.14 (early), 0.64 (middle), and 0.33 (late) for children.

### Association between health utilities and demographic/medical characteristics among adult participants

Using beta regression among adults aged ≥16 years, there was no evidence that health utilities differed by age, sex, education, or COVID-19 syndromes reported during infection (any symptoms, CLI, ILI, upper respiratory, lower respiratory, neurological, constitutional, moderate; Figure 3 and Supplementary Table 3). Results for crude and age-adjusted models for the full list of demographics/medical characteristics analyzed are presented as supplemental material (Supplemental Table 4).

**Figure 3.**
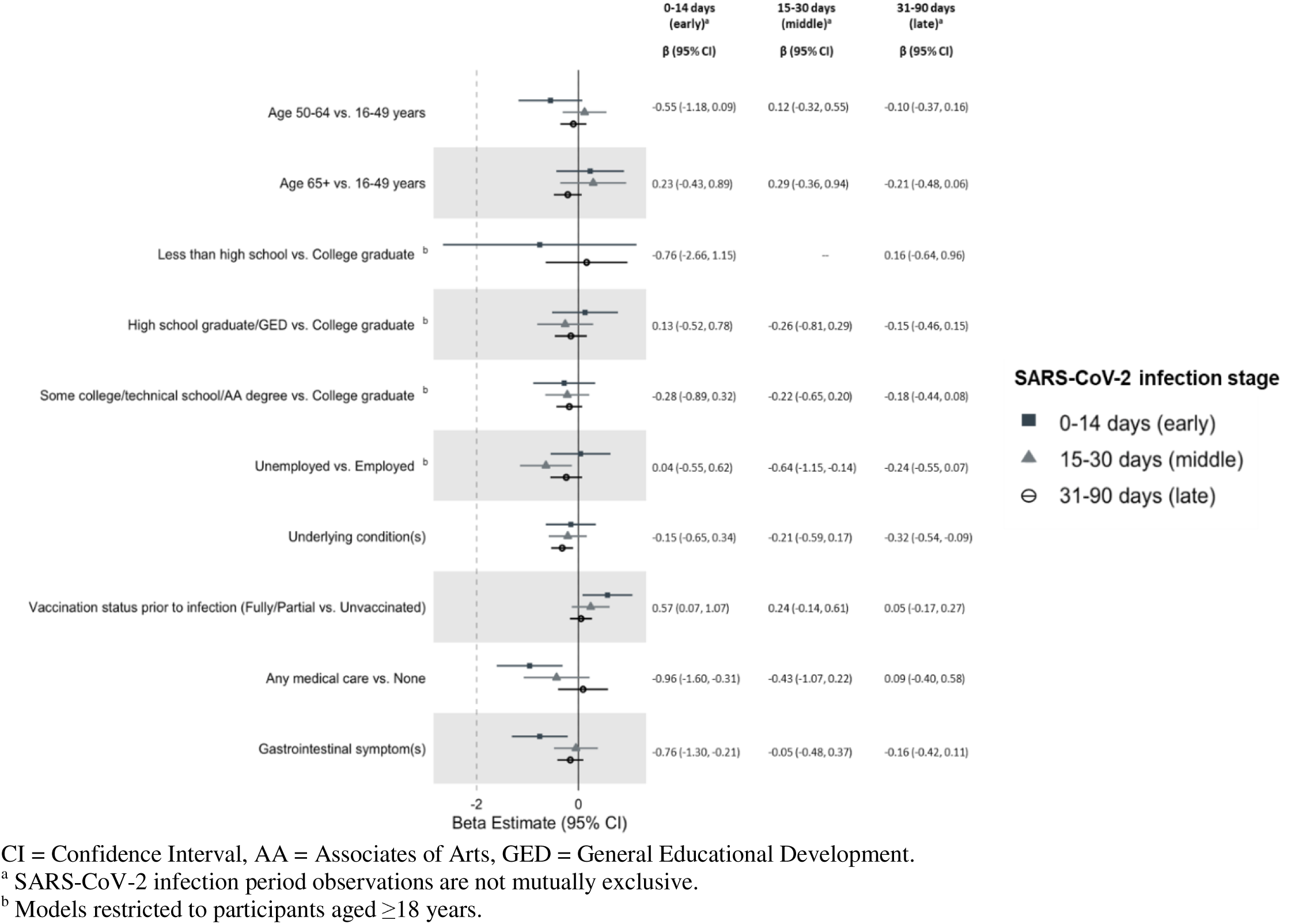
Age-adjusted associations between health utility and demographic/medical characteristics among adults aged ≥ 16 years with SARS-CoV-2 infection, by infection period. SARS-CoV-2 infection periods were defined by the time of the EQ-5D-3L survey after symptom onset/positive test.

Among adults who completed an EQ-5D-3L survey during the early infection period (0-14 days after symptom onset or positive test), participants who were fully or partially vaccinated prior to infection had a higher health utility (0.57 higher among those vaccinated) compared to those who were unvaccinated (age-adjusted Beta: 0.57 95% CI: 0.07, 1.07). Participants who self-reported receiving any type of medical care had significantly lower health utility compared to those who did not report seeking medical care (age-adjusted Beta: −0.96, 95% CI: −1.60, - 0.31). Participants with gastrointestinal symptoms also had significantly lower health utility (age-adjusted Beta: - 0.76 95% CI: −1.30, −0.21) (Figure 3).

Among adults who completed an EQ-5D-3L survey during the middle infection period, unemployed participants had lower health utility than employed participants (age-adjusted Beta: −0.64, 95% CI: −1.15, −0.14) (Figure 3).

Among adults who completed an EQ-5D-3L survey during the late infection period, participants with reported underlying conditions had lower health utility than those who did not (age-adjusted Beta: −0.32, 95% CI: - 0.54, −0.09) (Figure 3).

### Change in health state from enrollment

Among participants who completed the EQ-5D-3L survey both at enrollment and during the early infection period (n=61 out of 70 adults; n=22 out of 24 children), 36% of adults and 64% of children reported no change in their health from enrollment to early infection, 16% of adults and 5% of children reported improved health, and 38% of adults and 23% of children reported worsened health (Figure 4A). Worsened health was most frequently reported for domains of pain/discomfort (adults: 33%; children: 18%) and usual activities (adults: 30%; children 9%; Figure 4B).

**Figure 4.**
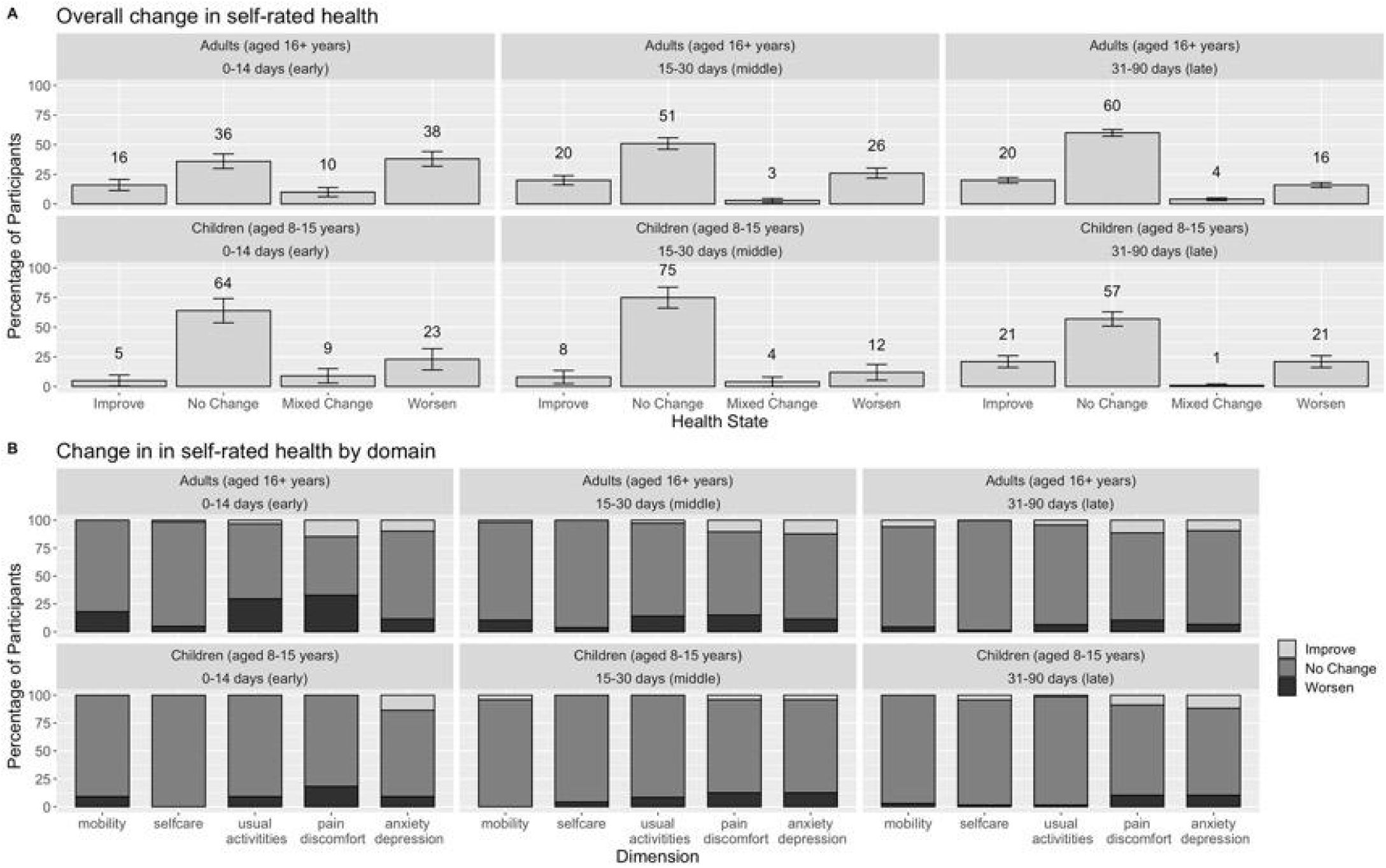
Change in self-rated health status (overall [A] and by domain [B]) from enrollment among participants with SARS-CoV-2 infection, by infection period among adults aged ≥16 years and children aged 8-15 years. Among participants with completed EQ-5D-3L surveys at enrollment and ≤90 days after symptom onset/positive test. SARS-CoV-2 infection periods were defined by the time of the EQ-5D-3L survey after symptom onset/positive test.

Among participants with EQ-5D-3L data at both enrollment and the middle infection period (n=106 out of 117 adults; n=24 out of 27 children), 51% of adults and 75% of children reported no change in their health during infection, 20% of adults and 8% of children reported improved health, and 26% of adults and 12% of children reported worsened health (Figure 4A).

Among participants with EQ-5D-3L survey at both enrollment and the late infection period (n=298 out of 351 adults; n=68 out of 73 children), 60% of adults and 57% of children reported no change in their health during infection, 20% of adults and 21% of children reported improved health, and 16% of adults and 21% of children reported worsened health (Figure 4A).

### Change in health state from enrollment by demographic/medical characteristics among adults

Among demographic/medical characteristics that showed significant associations with health utilities in the multivariate regression model with sufficient sample size for stratification in each infection period (n>5; Supplemental Table 5), having gotten vaccinated against COVID-19 prior to infection (fully or partially) was significantly associated with less negative health changes reported during early infection period; 54% of participants (n=14 out of 26) who were unvaccinated prior to their infection reported health was worsened vs. 26% (n=9 out of 35) of vaccinated participants (p<0.01).

## Discussion

Using data from three prospective cohorts in the U.S., health utilities were calculated for three post-SARS-CoV-2 infection periods ≤90 days post-symptom onset/positive test. Compared with reported health at enrollment, 38% of adults aged ≥16 years and 23% of children aged 5-18 years with SARS-CoV-2 experienced worsened health in the first 2 weeks after symptom onset or first positive test. Worsened health was most commonly due to pain/discomfort for all participants and having a harder time performing usual activities among adults. There was evidence that adults who were vaccinated with at least one dose against SARS-CoV-2 had improved health during the first 2 weeks of infection compared with those who were unvaccinated.

Most of the infections in the cohorts were symptomatic but mild, with only 8% reporting seeking medical care during their illness, and most reporting no problems with their health during all infection periods. This outcome may be partially explained by the demographic composition of our sample. Despite over half the sample having underlying conditions (53.7%), the minority of participants did not have other risk factors for severe COVID-19 and hospitalization (aged ≥65 years, certain races and ethnicities, and being unvaccinated against SARS-CoV-2).[31, 32] However, there was a sizeable proportion of adult participants who reported a worsening in health during SARS-CoV-2 infection, most commonly due to pain/discomfort. Problems were more frequently reported during the first 2 weeks after infection compared to later periods of infection. Our findings were consistent with other EQ-5D studies of patients with mild to severe COVID-19, where problems due to pain/discomfort during illness compared to the other domains (mobility, self-care, usual activities, anxiety/depression) are most frequently reported.[12, 33, 34] “COVID-pain” has been described by previous studies as a vast spectrum of clinical manifestations of the COVID-19 illness.^31^ The characterization includes myalgia, joint pain, sore throat, abdominal pain, chest pain, and headache. These are common symptoms associated with acute infection and may be similar to what was observed in these cohorts.[35, 36]

Consistent with existing literature exploring impact of SARS-CoV-2 infection among children, health remained largely unaffected among children in our study. Another EQ-5D study among children with SARS-CoV-2 infection found that most children with persistent symptoms reported mild or no impairment in their quality of life, including school function, 3-6 months after infection.[37] In this study, the proportion of children with worsened health over time was approximately half of the proportion of adults who reported worsened health. While children and adults have similar incidence rates of SARS-CoV-2 infection,^32^ children tend to have milder symptoms or be asymptomatic.[38, 39] Differences in immune response, tobacco use, alcohol consumption, and comorbidities have been proposed explanations for illness severity gaps between adults and children.[40, 41]

Despite overall high health utilities reported among these cohorts, there was variation when stratified by demographic and medical characteristics at different post-infection periods. Vaccination against SARS-CoV-2 had a positive effect on health utility among adults during the first 14 days after symptom onset/positive test. Our findings are consistent with other studies conducted among those with SARS-CoV-2 infection, which observed increased health utility and VAS scores among participants vaccinated against SARS-CoV-2 compared to those unvaccinated.[42, 43] Vaccinated individuals have also been shown to have less of a health decline, via health utility and VAS scores, from enrollment to up to four weeks post-positive RT-PCR test compared to unvaccinated counterparts.[42] Similarly, vaccinated participants were less likely to report worsened health from enrollment to £14 days post-symptom onset/positive test.

Adults who sought medical care during their illness and those with gastrointestinal symptoms had significantly worse self-reported health during the first two weeks of infection. These findings support one another, given that individuals who seek medical care generally have more severe symptoms.[44] However, these risk factors were transient and not significantly related to worse health in later weeks after infection, as we defined them in this study at 31-90 days. Unemployed adults had lower health utility in the subsequent two weeks after infection. Other studies have reported increased illness severity and ICU admittance among unemployed adults with COVID-19.[45, 46] Not only do unemployed individuals have limited access to medical care/resources,[47] but they also have been suggested to be older in age, a main risk factor for severe illness.[45] Lower health utilities in the months after infection, in this study, were only associated with having underlying conditions. Other studies have also observed that having underlying conditions can increase the risk of lingering symptoms or developing post/long COVID.[48]

### Strengths and Limitations

This study combined data from three large cohorts from different U.S. locations followed during similar timeline and with comparable methods, including standardized implementation of the EQ-5D-3L survey to measure health utility and the VAS to measure self-reported health. A further strength of the study was the large number of SARS-CoV-2 infections that were detected. Because we collected EQ-5D-3L data at multiple time points, we were able to observe changes in self-rated health over time. However, at least four limitations should be considered when interpreting our findings. First, even with a large number of SARS-CoV-2 infections, the cohorts had limited diversity in some of the demographic and medical characteristics, which limited our ability to fully adjust for potential confounding and participants may not be representative of the general population in the U.S. With a relatively young and healthy sample, our estimates likely underestimate health-related quality of life impact of SARS-CoV-2 infection. Relatedly, we were only able to present limited stratified results for participants aged 8-15 years due to small numbers. Second, we were unable to conduct time series analysis because few participants completed more than one EQ-5D-3L survey after illness or infection onset. Third, the EQ-5D-3L surveys in the C-HEaRT and SEARCh cohorts conducted after an infection asked about health during the illness and, thus, may not be representative of convalescent health. Lastly, results regarding vaccination did not take into account time since vaccination, an important determinant of effectiveness.

### Conclusion

Among participants infected with SARS-CoV-2 across three cohorts followed from August 2020 to July 2022, self-rated health at various times during 90 days after infection onset infection was lower in those who sought medical care, had gastrointestinal symptoms, were unemployed, and had underlying health conditions. Vaccinated participants were less likely to experience a health decline during the first two weeks following infection than unvaccinated participants. Overall, health-related quality of life measured by health utilities, VAS scores, and Paretian Classification of Health Change was largely unaffected by non-hospitalized SARS-CoV-2 infection. Although health utilities were impacted most during the first 2 weeks of infection, population-level impacts are high given the high infection rates during peak circulation of SARS-CoV-2 viruses. Data from this study may support future evaluations of the full burden of SARS-CoV-2 infection and benefits of interventions, such as vaccination.

## Data Availability

Data in the present work may be available upon reasonable request to the authors.

## Acknowledgments

We thank the following for their contributions to the study: Carrie Reed, Natalie Thornburg, and Alicia Fry at the Centers for Disease Control and Prevention; Priyam Thind and Maria Castro at Columbia University; Edward Belongia, Jennifer King, Roxy Eibergen, Carla Rottscheit, Erik Kronholm, and Kayla Hanson at Marshfield Clinic Research Institute.

## References

1. Centers of Disease Control and Prevention. CDC Museum COVID-19 Timeline. 2023. https://www.cdc.gov/museum/timeline/covid19.html#:∼:text=January%2020%2C%202020,respond%20to%20the%20emerging%20outbreak. Accessed Aug 6, 2024.

2. Centers of Disease Control and Prevention. COVID Data Tracker. 2024. https://covid.cdc.gov/covid-data-tracker/#trends_totaldeaths_select_00. Accessed Aug 6, 2024.

3. Edouard Mathieu HR, Lucas Rodés-Guirao, Cameron Appel, Charlie Giattino, Joe Hasell, Bobbie Macdonald, Saloni Dattani, Diana Beltekian, Esteban Ortiz-Ospina and Max Roser. Coronavirus (COVID-19) Hospitalizations. 2020. https://ourworldindata.org/covid-hospitalizations. Accessed

4. Kaye AD, Okeagu CN, Pham AD, Silva RA, Hurley JJ, Arron BL, Sarfraz N, Lee HN, Ghali GE, Gamble JW et al. Economic impact of COVID-19 pandemic on healthcare facilities and systems: International perspectives. Best Practice & Research Clinical Anaesthesiology 2021, 35(3):293–306. DOI: 10.1016/j.bpa.2020.11.009

5. Malik P, Patel K, Pinto C, Jaiswal R, Tirupathi R, Pillai S, Patel U. Post-acute COVID-19 syndrome (PCS) and health-related quality of life (HRQoL)-A systematic review and meta-analysis. J Med Virol 2022, 94(1):253–262. DOI: 10.1002/jmv.27309

6. Menges D, Ballouz T, Anagnostopoulos A, Aschmann HE, Domenghino A, Fehr JS, Puhan MA. Burden of post-COVID-19 syndrome and implications for healthcare service planning: A population-based cohort study. PLoS One 2021, 16(7):e0254523. DOI: 10.1371/journal.pone.0254523

7. Hay JW, Gong CL, Jiao X, Zawadzki NK, Zawadzki RS, Pickard AS, Xie F, Crawford SA, Gu NY. A US Population Health Survey on the Impact of COVID-19 Using the EQ-5D-5L. Journal of General Internal Medicine 2021, 36(5):1292–1301. DOI: 10.1007/s11606-021-06674-z

8. Centers of Disease Control and Prevention. Long COVID Basics. 2024. https://www.cdc.gov/covid/long-term-effects/?CDC_AAref_Val=https://www.cdc.gov/coronavirus/2019-ncov/long-term-effects/index.html. Accessed Jun 3, 2025.

9. World Health Organization. Post COVID-19 condition (Long COVID). 2022. https://www.who.int/europe/news-room/fact-sheets/item/post-covid-19-condition. Accessed Jun 6, 2025.

10. Devlin N, Parkin D, Janssen B: Methods for analysing and reporting EQ-5D data: Springer Nature; 2020.

11. Cazé AB, Cerqueira-Silva T, Bomfim AP, de Souza GL, Azevedo AC, Brasil MQ, Santos NR, Khouri R, Dan J, Bandeira AC et al. Prevalence and risk factors for long COVID after mild disease: A cohort study with a symptomatic control group. J Glob Health 2023, 13:06015. DOI: 10.7189/jogh.13.06015

12. Smith P, De Pauw R, Van Cauteren D, Demarest S, Drieskens S, Cornelissen L, Devleesschauwer B, De Ridder K, Charafeddine R. Post COVID-19 condition and health-related quality of life: a longitudinal cohort study in the Belgian adult population. BMC Public Health 2023, 23(1):1433. DOI: 10.1186/s12889-023-16336-w

13. Alacevich C, Thalmann I, Nicodemo C, de Lusignan S, Petrou S. Symptomatic SARS-CoV-2 Episodes and Health-Related Quality of Life. Appl Health Econ Health Policy 2023, 21(5):761–771. DOI: 10.1007/s40258-023-00810-y

14. Domazet Bugarin J, Saric L, Delic N, Dosenovic S, Ilic D, Saric I, Stipic SS, Duplancic B. Health-Related Quality of Life of COVID-19 Survivors Treated in Intensive Care Unit-Prospective Observational Study. J Intensive Care Med 2023, 38(8):710–716. DOI: 10.1177/08850666231158547

15. Herrmann J, Müller K, Notz Q, Hübsch M, Haas K, Horn A, Schmidt J, Heuschmann P, Maschmann J, Frosch M et al. Prospective single-center study of health-related quality of life after COVID-19 in ICU and non-ICU patients. Sci Rep 2023, 13(1):6785. DOI: 10.1038/s41598-023-33783-y

16. Mitrović-Ajtić O, Stanisavljević D, Miljatović S, Dragojević T, Živković E, Šabanović M, Čokić VP. Quality of Life in Post-COVID-19 Patients after Hospitalization. Healthcare (Basel) 2022, 10(9). DOI: 10.3390/healthcare10091666

17. Sumner KM, Karron RA, Stockwell MS, Dawood FS, Stanford JB, Mellis A, Hacker E, Thind P, Castro MJE, Harris JP et al. Impact of Age and Symptom Development on SARS-CoV-2 Transmission in Households With Children-Maryland, New York, and Utah, August 2020-October 2021. Open Forum Infect Dis 2022, 9(8):ofac390. DOI: 10.1093/ofid/ofac390

18. McLean HQ, McClure DL, King JP, Meece JK, Pattinson D, Neumann G, Kawaoka Y, Rolfes MA, Belongia EA. mRNA COVID-19 vaccine effectiveness against SARS-CoV-2 infection in a prospective community cohort, rural Wisconsin, November 2020 to December 2021. Influenza Other Respir Viruses 2022, 16(4):607–612. DOI: 10.1111/irv.12970

19. Devlin NJ, Brooks R. EQ-5D and the EuroQol Group: Past, Present and Future. Appl Health Econ Health Policy 2017, 15(2):127–137. DOI: 10.1007/s40258-017-0310-5

20. Shaw JW, Johnson JA, Coons SJ. US valuation of the EQ-5D health states: development and testing of the D1 valuation model. Med Care 2005, 43(3):203–220. DOI: 10.1097/00005650-200503000-00003

21. Janssen MF, Bonsel GJ, Luo N. Is EQ-5D-5L Better Than EQ-5D-3L? A Head-to-Head Comparison of Descriptive Systems and Value Sets from Seven Countries. PharmacoEconomics 2018, 36(6):675–697. DOI: 10.1007/s40273-018-0623-8

22. Gerlinger C, Bamber L, Leverkus F, Schwenke C, Haberland C, Schmidt G, Endrikat J. Comparing the EQ-5D-5L utility index based on value sets of different countries: impact on the interpretation of clinical study results. BMC Research Notes 2019, 12(1):18. DOI: 10.1186/s13104-019-4067-9

23. Ramos-Goñi JM, Oppe M, Estévez-Carrillo A, Rivero-Arias O, Wolfgang G, Simone K, Kristina L, Valentina R. Accounting for Unobservable Preference Heterogeneity and Evaluating Alternative Anchoring Approaches to Estimate Country-Specific EQ-5D-Y Value Sets: A Case Study Using Spanish Preference Data. Value in Health 2022, 25(5):835–843. DOI: 10.1016/j.jval.2021.10.013

24. Hoogeveen MJ, Hoogeveen EK. Comparable seasonal pattern for COVID-19 and flu-like illnesses. One Health 2021, 13:100277. DOI: 10.1016/j.onehlt.2021.100277

25. Bozio CH, Grannis SJ, Naleway AL, Ong TC, Butterfield KA, DeSilva MB, Natarajan K, Yang D-H, Rao S, Klein NP. Laboratory-confirmed COVID-19 among adults hospitalized with COVID-19–like illness with infection-induced or mRNA vaccine-induced SARS-CoV-2 immunity—nine states, January–September 2021. Morbidity and Mortality Weekly Report 2021, 70(44):1539. DOI: 10.15585/mmwr.mm7044e1

26. McLean HQ, Grijalva CG, Hanson KE, Zhu Y, Deyoe JE, Meece JK, Halasa NB, Chappell JD, Mellis AM, Reed C. Household transmission and clinical features of SARS-CoV-2 infections. Pediatrics 2022, 149(3):e2021054178. DOI:

27. R Core Team. R: A Language and Environment for Statistical Computing. 2022. https://www.R-project.org/. Accessed

28. Nijjar FMaJS: eq5d: Methods for Analysing ‘EQ-5D’ Data and Calculating ‘EQ-5D’ Index Scores. In.; 2023: R package version 0.15.10}.

29. Reckers-Droog V, Karimi M, Lipman S, Verstraete J. Why Do Adults Value EQ-5D-Y-3L Health States Differently for Themselves Than for Children and Adolescents: A Think-Aloud Study. Value in Health 2022, 25(7):1174–1184. DOI: 10.1016/j.jval.2021.12.014

30. Moberg C, Alderling M, Meding B. Hand eczema and quality of life: a population-based study. Br J Dermatol 2009, 161(2):397–403. DOI: 10.1111/j.1365-2133.2009.09099.x

31. Zhang JJ, Dong X, Liu GH, Gao YD. Risk and Protective Factors for COVID-19 Morbidity, Severity, and Mortality. Clin Rev Allergy Immunol 2023, 64(1):90–107. DOI: 10.1007/s12016-022-08921-5

32. Tenforde MW, Self WH, Adams K, Gaglani M, Ginde AA, McNeal T, Ghamande S, Douin DJ, Talbot HK, Casey JD et al. Association Between mRNA Vaccination and COVID-19 Hospitalization and Disease Severity. JAMA 2021, 326(20):2043–2054. DOI: 10.1001/jama.2021.19499

33. Román-Montes CM, Flores-Soto Y, Guaracha-Basañez GA, Tamez-Torres KM, Sifuentes-Osornio J, González-Lara MF, de León AP. Post-COVID-19 syndrome and quality of life impairment in severe COVID-19 Mexican patients. Front Public Health 2023, 11:1155951. DOI: 10.3389/fpubh.2023.1155951

34. Moens M, Duarte RV, De Smedt A, Putman K, Callens J, Billot M, Roulaud M, Rigoard P, Goudman L. Health-related quality of life in persons post-COVID-19 infection in comparison to normative controls and chronic pain patients. Front Public Health 2022, 10:991572. DOI: 10.3389/fpubh.2022.991572

35. Cascella M, Del Gaudio A, Vittori A, Bimonte S, Del Prete P, Forte CA, Cuomo A, De Blasio E. COVID-Pain: Acute and Late-Onset Painful Clinical Manifestations in COVID-19 - Molecular Mechanisms and Research Perspectives. J Pain Res 2021, 14:2403–2412. DOI: 10.2147/jpr.S313978

36. Shanthanna H, Nelson AM, Kissoon N, Narouze S. The COVID-19 pandemic and its consequences for chronic pain: a narrative review. Anaesthesia 2022, 77(9):1039–1050. DOI: 10.1111/anae.15801

37. Franco JVA, Garegnani LI, Oltra GV, Metzendorf M-I, Trivisonno LF, Sgarbossa N, Ducks D, Heldt K, Mumm R, Barnes B et al. Short and Long-Term Wellbeing of Children following SARS-CoV-2 Infection: A Systematic Review. International Journal of Environmental Research and Public Health 2022, 19(21):14392. DOI:

38. Mehta NS, Mytton OT, Mullins EWS, Fowler TA, Falconer CL, Murphy OB, Langenberg C, Jayatunga WJP, Eddy DH, Nguyen-Van-Tam JS. SARS-CoV-2 (COVID-19): What Do We Know About Children? A Systematic Review. Clinical Infectious Diseases 2020, 71(9):2469–2479. DOI: 10.1093/cid/ciaa556

39. Dawood FS, Porucznik CA, Veguilla V, Stanford JB, Duque J, Rolfes MA, Dixon A, Thind P, Hacker E, Castro MJE, et al. Incidence Rates, Household Infection Risk, and Clinical Characteristics of SARS-CoV-2 Infection Among Children and Adults in Utah and New York City, New York. JAMA Pediatr 2022, 176(1):59–67. DOI: 10.1001/jamapediatrics.2021.4217

40. Chou J, Thomas PG, Randolph AG. Immunology of SARS-CoV-2 infection in children. Nature Immunology 2022, 23(2):177–185. DOI: 10.1038/s41590-021-01123-9

41. Morrell ED, Mikacenic C. Differences between Children and Adults with COVID-19: It’s Right under Our Nose. American Journal of Respiratory Cell and Molecular Biology 2022, 66(2):122–123. DOI: 10.1165/rcmb.2021-0455ED

42. Fusco MD, Sun X, Moran MM, Coetzer H, Zamparo JM, Puzniak L, Alvarez MB, Tabak YP, Cappelleri JC. Impact of COVID-19 and Effects of Vaccination with BNT162b2 on Patient-Reported Health-Related Quality of Life, Symptoms, and Work Productivity Among US Adult Outpatients with SARS-CoV-2. medRxiv 2022, 10.1101/2022.08.31.22279264:2022.2008.2031.22279264. DOI: 10.1101/2022.08.31.22279264

43. Kuodi P, Gorelik Y, Zayyad H, Wertheim O, Beiruti Wiegler K, Abu Jabal K, Dror AA, Elsinga J, Nazzal S, Glikman D et al. Association between BNT162b2 vaccination and health-related quality of life up to 18 months post-SARS-CoV-2 infection in Israel. Scientific Reports 2023, 13(1):15801. DOI: 10.1038/s41598-023-43058-1

44. Kim L, Garg S, O’Halloran A, Whitaker M, Pham H, Anderson EJ, Armistead I, Bennett NM, Billing L, Como-Sabetti K et al. Risk Factors for Intensive Care Unit Admission and In-hospital Mortality Among Hospitalized Adults Identified through the US Coronavirus Disease 2019 (COVID-19)-Associated Hospitalization Surveillance Network (COVID-NET). Clin Infect Dis 2021, 72(9):e206–e214. DOI: 10.1093/cid/ciaa1012

45. Alharbi A-HM, Rabbani SI, Halim Mohamed AA, Almushayti BK, Aldhwayan NI, Almohaimeed AT, Alharbi AA, Alharbi NS, Asdaq SMB, Alamri AS. Analysis of potential risk factors associated with COVID-19 and hospitalization. Frontiers in public health 2022, 10:921953. DOI:

46. Núñez Cortés RI, Ortega Palavecinos M, Soto Carmona CA, Torres Gangas P, Rivero C, Paz M, Torres Castro RH. Social determinants of health associated with severity and mortality in patients with COVID-19. 2021. DOI:

47. Berkowitz SA, Basu S. Unemployment Insurance, Health-Related Social Needs, Health Care Access, and Mental Health During the COVID-19 Pandemic. JAMA Internal Medicine 2021, 181(5):699–702. DOI: 10.1001/jamainternmed.2020.7048

48. Centers for Disease Control and Prevention. Long COVID or Post-COVID Conditions. 2022. https://www.cdc.gov/covid/long-term-effects/?CDC_AAref_Val=https://www.cdc.gov/coronavirus/2019-ncov/long-term-effects/index.html. Accessed Jun 3, 2025.

