## Supplemental information for "Estimating Impact of SARS-CoV-2 Infection on Health-Related Quality of Life among Persons Aged 8 years and Older, August 2020-July 2022"

Supplemental Table 1. Characteristics of participants aged ≥8 years with SARS-CoV-2 infection with and without EQ-5D survey data within 90 days after symptom onset/positive test from three prospective, longitudinal cohorts in the United States from August 2020-July 2022

| | Periods after onset/ positive test No.(%)[a,b] | | | | |
|---|---|---|---|---|---|
| | Participants with SARS-CoV-2 infection[c] | Participants with SARS-CoV-2 infection without EQ-5D data within 90 days after infection | Participants with SARS-CoV-2 infection with EQ-5D data during 0-14 days after infection (early) | Participants with SARS-CoV-2 infection with EQ-5D data during 15-30 days after infection (middle) | Participants with SARS-CoV-2 infection with EQ-5D data during 31-90 days after infection (late) |
| No. | (n=763) | (n=188) | (n=94) | (n=144) | (n=424) |
| Location (Cohort) | | | | | |
| Maryland (SEARCh) | 24 (3.1) | 7 (3.7) | 6 (6.4) | 11 (7.6) | 2 (0.5) |
| New York (C-HEaRT) | 21 (2.8) | 8 (4.3) | 6 (6.4) | 7 (4.9) | 1 (0.2) |
| Utah (C-HEaRT) | 66 (8.7) | 24 (12.8) | 22 (23.4) | 15 (10.4) | 14 (3.3) |
| Wisconsin (PACC) | 652 (85.5) | 149 (79.3) | 60 (63.8) | 111 (77.1) | 407 (96.0) |
| Sex | | | | | |
| Female | 433 (56.7) | 102 (54.3) | 55 (58.5) | 88 (61.1) | 242 (57.1) |
| Male | 330 (43.3) | 86 (45.7) | 39 (41.5) | 56 (38.9) | 182 (42.9) |
| Age (in years) | | | | | |
| Median (IQR) | 40.00 (24.00, 59.00) | 40.00 (18.75, 61.00) | 34.50 (15.25, 45.75) | 38.00 (25.00, 52.00) | 41.50 (27.00, 59.00) |
| 8-15 | 147 (19.3) | 41 (21.8) | 24 (25.5) | 27 (18.8) | 73 (17.2) |
| 16-49 | 338 (44.3) | 72 (38.3) | 48 (51.1) | 76 (52.8) | 189 (44.6) |
| 50-64 | 140 (18.3) | 31 (16.5) | 11 (11.7) | 30 (20.8) | 84 (19.8) |
| 65+ | 138 (18.1) | 44 (23.4) | 11 (11.7) | 11 (7.6) | 78 (18.4) |
| Race/Ethnicity | | | | | |
| Asian, non-Hispanic | 3 (0.4) | 0 (0.0) | 1 (1.1) | 2 (1.4) | 0 (0.0) |
| Black, non-Hispanic | 4 (0.5) | 1 (0.5) | 0 (0.0) | 1 (0.7) | 3 (0.7) |
| White, non-Hispanic | 705 (92.4) | 165 (87.8) | 84 (89.4) | 130 (90.3) | 409 (96.5) |
| Multiracial, non-Hispanic | 8 (1.0) | 2 (1.1) | 2 (2.1) | 2 (1.4) | 3 (0.7) |
| Hispanic | 42 (5.5) | 19 (10.1) | 7 (7.4) | 9 (6.2) | 9 (2.1) |
| Unknown race | 1 (0.1) | 1 (0.5) | 0 (0.0) | 0 (0.0) | 0 (0.0) |
| Education | | | | | |
| Age <18 years | 169 (22.1) | 46 (24.5) | 28 (29.8) | 30 (20.8) | 87 (20.5) |
| Less than high school | 250 (32.8) | 64 (34.0) | 32 (34.0) | 49 (34.0) | 133 (31.4) |
| High school graduate/GED | 14 (1.8) | 6 (3.2) | 1 (1.1) | 0 (0.0) | 7 (1.7) |
| Some college/technical school/AA degree | 130 (17.0) | 34 (18.1) | 16 (17.0) | 19 (13.2) | 72 (17.0) |
| College graduate | 195 (25.6) | 37 (19.7) | 17 (18.1) | 45 (31.2) | 122 (28.8) |
| Employment status | | | | | |
| Age <18 years | 169 (22.1) | 46 (24.5) | 28 (29.8) | 30 (20.8) | 87 (20.5) |
| Unemployed | 177 (23.2) | 53 (28.2) | 22 (23.4) | 27 (18.8) | 89 (21.0) |
| Employed | 414 (54.3) | 89 (47.3) | 44 (46.8) | 86 (59.7) | 246 (58.0) |
| Underlying condition(s) [d] | | | | | |
| Yes | 353 (46.3) | 87 (46.3) | 46 (48.9) | 60 (41.7) | 196 (46.2) |
| No | 410 (53.7) | 101 (53.7) | 48 (51.1) | 84 (58.3) | 228 (53.8) |
| COVID-19 vaccination status prior to infection[e] | | | | | |
| Unvaccinated | 352 (46.1) | 86 (45.7) | 51 (54.3) | 77 (53.5) | 184 (43.4) |
| Partially vaccinated | 12 (1.6) | 1 (0.5) | 1 (1.1) | 3 (2.1) | 7 (1.7) |
| Fully vaccinated | 388 (50.9) | 90 (47.9) | 42 (44.7) | 64 (44.4) | 233 (55.0) |
| Self-reported symptom(s) ≤30 days post-onset/positive test[f] | | | | | |
| Yes | 507 (66.4) | 71 (37.8) | 78 (83.0) | 122 (84.7) | 297 (70.0) |
| No | 92 (12.1) | 31 (16.5) | 16 (17.0) | 22 (15.3) | 29 (6.8) |
| Days between symptom onset/positive test and EQ-5D-3L/ VAS completion | | | | | |
| Mean (SD) | 37.85 (22.22) | — | 8.34 (3.94) | 23.40 (4.96) | 52.45 (17.44) |
| Median (IQR) | 34.00 (21.00, 53.50) | — | 8.50 (6.00, 12.00) | 23.00 (20.00, 28.00) | 50.00 (36.00, 66.00) |

[a] Percentage may not sum to 100 due to missing values
[b] Unless specified, characteristics represent enrollment status
[c] Represents unique number of participants. Participants may be in multiple infection periods
[d] Self-reported at least one of the following: obesity, cancer, kidney disease, COPD, hypertension, immunocompromised, liver disease, heart disease, mental conditions, and diabetes
[e] Partially vaccinated: received 1 dose of 2 dose SARS-CoV-2 vaccine; Fully vaccinated: received 2 doses of 2 dose SARS-CoV-2 vaccine or 1 dose of 1 dose SARS-CoV-2 vaccine
[f] Self-reported at least one of the following: fever, cough, loss/change in taste/smell, sore throat, muscle/body aches, shortness of breath, diarrhea, fatigue, headache, nasal congestion, vomiting

Supplemental Table 2. Characteristics of adult participants aged ≥8 years with SARS-CoV-2 infection and EQ-5D-3L survey data within 90 days after symptom onset/positive test from three prospective, longitudinal cohorts in the United States from August 2020-July 2022

| | Cohort No.(%)[a,b,c] | | |
|---|---|---|---|
| Characteristic | C-HEaRT (n=55) | PACC (n=503) | SEARCh (n=17) |
| Sex | | | |
| Female | 32 (58.2) | 289 (57.5) | 10 (58.8) |
| Male | 23 (41.8) | 214 (42.5) | 7 (41.2) |
| Age (in years) | | | |
| Median (IQR) | 33.00 (16.5, 41.0) | 42.00 (27.0, 60.0) | 35.00 (32.00, 38.00) |
| 08-15 | 12 (21.8) | 92 (18.3) | 2 (11.8) |
| 16-49 | 41 (74.5) | 210 (41.7) | 15 (88.2) |
| 50-64 | 2 (3.6) | 107 (21.3) | 0 (0.0) |
| 65+ | 0 (0.0) | 94 (18.7) | 0 (0.0) |
| Race/Ethnicity | | | |
| Asian, non-Hispanic | 0 (0.0) | 1 (0.2) | 2 (11.8) |
| Black, non-Hispanic | 1 (1.8) | 1 (0.2) | 1 (5.9) |
| White, non-Hispanic | 41 (74.5) | 488 (97.0) | 11 (64.7) |
| Multiracial, non-Hispanic | 0 (0.0) | 3 (0.6) | 3 (17.6) |
| Hispanic | 13 (23.6) | 10 (2.0) | 0 (0.0) |
| Education | | | |
| Age <18 years | 16 (29.1) | 105 (20.9) | 2 (11.8) |
| Less than high school (adult) | 1 (1.8) | 7 ( 1.4) | 0 (0.0) |
| High school grad/GED | 6 (10.9) | 87 (17.3) | 3 (17.6) |
| Some college/technical school/AA degree | 7 (12.7) | 149 (29.6) | 2 (11.8) |
| College grad | 25 (45.5) | 151 (30.0) | 10 (58.8) |
| Employment Status | | | |
| Age <18 years | 16 (29.1) | 105 (20.9) | 2 (11.8) |
| Employed | 28 (50.9) | 284 (56.5) | 13 (76.5) |
| Unemployed | 11 (20.0) | 111 (22.1) | 2 (11.8) |
| Any underlying condition(s)[d] | | | |
| Yes | 15 (27.3) | 241 (47.9) | 10 (58.8) |
| No | 40 (72.7) | 262 (52.1) | 7 (41.2) |
| Sought medical care[e] | | | |
| Yes, Outpatient care | 9 (16.4) | 0 ( 0.0) | 11 (64.7) |
| Yes, unknown medical care location | 1 (1.8) | 19 (3.8) | 0 (0.0) |
| No | 45 (81.8) | 390 (77.5) | 6 (35.3) |
| COVID-19 vaccination status prior to infection | | | |
| Unvaccinated | 42 (76.4) | 212 (42.1) | 14 (82.4) |
| Partially vaccinated | 2 ( 3.6) | 7 ( 1.4) | 1 ( 5.9) |
| Fully vaccinated | 11 (20.0) | 284 (56.5) | 2 (11.8) |
| Self-reported symptom(s)[e,g] | | | |
| Yes | 47 (85.5) | 360 (71.6) | 16 (94.1) |
| No | 8 (14.5) | 49 (9.7) | 1 (5.9) |

[a] Percentage may not sum to 100 due to missing values
[b] Unless specified, characteristics represent status at enrollment
[c] Represents unique number of participants.
[d] Self-reported at least one of the following: obesity, cancer, kidney disease, COPD, hypertension, immunocompromised, liver disease, heart disease, mental conditions, and diabetes
[e] Self-reported ≤30 days post-onset/positive test
[f] Partially vaccinated: received 1 dose of 2 dose SARS-CoV-2 vaccine; Fully vaccinated: received 2 doses of 2 dose SARS-CoV-2 vaccine or 1 dose of 1 dose SARS-CoV-2 vaccine
[g] Self-reported at least one of the following: fever, cough, loss/change in taste/smell, sore throat, muscle/body aches, shortness of breath, diarrhea, fatigue, headache, nasal congestion, vomiting

Supplemental Table 3. Descriptive statistics for health utilities among participants aged ≥8 years with SARS-CoV-2 infection period, by infection period stratified by age

| Participants ≥16 years[a] | | | | | |
|---|---|---|---|---|---|
| SARS CoV-2 infection period | No. | Mean (SD) | Median (IQR) | Bootstrapped 95% CI | Range |
| Overall | 538 | 0.90 (0.14) | 1.00 (0.83, 1.00) | 0.88,0.91 | 0.17,1.00 |
| 0-14 days (early) | 70 | 0.86 (0.16) | 0.83 (0.80, 1.00) | 0.82,0.89 | 0.22,1.00 |
| 15-30 days (middle) | 117 | 0.88 (0.17) | 1.00 (0.82, 1.00) | 0.85,0.9 | 0.17,1.00 |
| 31-90 days (late) | 351 | 0.91 (0.13) | 1.00 (0.83, 1.00) | 0.9,0.93 | 0.17,1.00 |
| Participants 8-15 years[b] | | | | | |
| SARS CoV-2 infection period | No. | Mean (SD) | Median (IQR) | Bootstrapped 95% CI | Range |
| Overall | 124 | 0.94 (0.12) | 1.00 (0.89, 1.00) | 0.91,0.96 | 0.12,1.00 |
| 0-14 days (early) | 24 | 0.93 (0.11) | 1.00 (0.89, 1.00) | 0.88,0.97 | 0.61,1.00 |
| 15-30 days (middle) | 27 | 0.90 (0.20) | 1.00 (0.89, 1.00) | 0.82,0.97 | 0.12,1.00 |
| 31-90 days (late) | 73 | 0.95 (0.08) | 1.00 (0.89, 1.00) | 0.93,0.97 | 0.71,1.00 |

SD = standard deviation; IQR = interquartile range; Cl = confidence interval

SARS-CoV-2 infection period = defined by the time of EQ-5D-3L survey after symptom onset/positive test

[a] US value sets for adults (aged ≥16 years) (Shaw JW, Johnson JA, Coons SJ. US valuation of the EQ-5D health states: development and testing of the D1 valuation model. *Med Care*. Mar 2005;43(3):203-20. doi:10.1097/00005650-200503000-00003)

[b] Value sets used to calculate health utilities derived from Spanish children (aged 8-15 years) onset/positive test (Ramos-Goñi JM, Oppe M, Estévez-Carrillo A, et al. Accounting for Unobservable Preference Heterogeneity and Evaluating Alternative Anchoring Approaches to Estimate Country-Specific EQ-5D-Y Value Sets: A Case Study Using Spanish Preference Data. *Value in Health*. 2022;25(5):835-843. doi:10.1016/j.jval.2021.10.013)

Supplemental Table 4. Associations between health utility and demographic/medical characteristics among adults aged ≥16 years

| | 0-14 days (early)[a] | | 15-30 days (middle)[a] | | 31-90 days (late)[a] | |
|---|---|---|---|---|---|---|
| Demographic/medical characteristics | Crude[b] β (95% CI) | Age-adjusted[b] β (95% CI) | Crude[b] β (95% CI) | Age-adjusted[b] β (95% CI) | Crude[b] β (95% CI) | Age-adjusted[b] β (95% CI) |
| Age 50-64 vs. 16-49 years | -0.55 (-1.18, 0.09) | -- | 0.12 (-0.32, 0.55) | -- | -0.10 (-0.37, 0.16) | -- |
| Age 65+ vs. 16-49 years | 0.23 (-0.43, 0.89) | -- | 0.29 (-0.36, 0.94) | -- | -0.21 (-0.48, 0.06) | -- |
| Sex (Male vs. Female) | 0.26 (-0.22, 0.74) | 0.26 (-0.23, 0.74) | 0.11 (-0.28, 0.50) | 0.08 (-0.31, 0.48) | -0.02 (-0.24, 0.20) | -0.01 (-0.23, 0.21) |
| Black, non-Hispanic vs. White, non-Hispanic[c] | -- | -- | **-2.73 (-4.61, -0.84)** | **-2.64 (-4.53, -0.75)** | 0.52 (-0.65, 1.69) | 0.49 (-0.68, 1.66) |
| Asian, non-Hispanic vs. White, non-Hispanic | -- | -- | 0.62 (-0.82, 2.06) | 0.67 (-0.77, 2.11) | -- | -- |
| Multiracial, non-Hispanic vs. White, non-Hispanic[c] | **-2.06 (-3.81, -0.31)** | **-2.08 (-3.86, -0.29)** | 0.62 (-1.41, 2.64) | 0.76 (-1.29, 2.80) | 0.52 (-0.91, 1.95) | 0.48 (-0.95, 1.91) |
| Hispanic vs. White, non-Hispanic | -0.20 (-1.01, 0.62) | -0.20 (-1.03, 0.63) | -0.37 (-1.20, 0.46) | -0.33 (-1.16, 0.51) | 0.20 (-0.57, 0.97) | 0.19 (-0.58, 0.96) |
| Less than high school vs. College graduate[d] | -0.70 (-2.60, 1.19) | -0.76 (-2.66, 1.15) | -- | -- | 0.06 (-0.72, 0.84) | 0.16 (-0.64, 0.96) |
| High school graduate/GED vs. College graduate[d] | 0.18 (-0.43, 0.78) | 0.13 (-0.52, 0.78) | -0.19 (-0.73, 0.36) | -0.26 (-0.81, 0.29) | -0.21 (-0.51, 0.08) | -0.15 (-0.46, 0.15) |
| Some college/technical school/AA degree vs. College graduate[d] | -0.24 (-0.82, 0.34) | -0.28 (-0.89, 0.32) | -0.15 (-0.57, 0.27) | -0.22 (-0.65, 0.20) | -0.21 (-0.46, 0.04) | -0.18 (-0.44, 0.08) |
| Unemployed vs. Employed[d] | 0.09 (-0.42, 0.60) | 0.04 (-0.55, 0.62) | -0.33 (-0.78, 0.11) | **-0.64 (-1.15, -0.14)** | -0.26 (-0.51, -0.01) | -0.24 (-0.55, 0.07) |
| Underlying condition(s) | -0.17 (-0.65, 0.31) | -0.24 (-0.75, 0.26) | -0.17 (-0.54, 0.20) | -0.21 (-0.59, 0.17) | **-0.33 (-0.55, -0.12)** | **-0.32 (-0.54, -0.09)** |
| Vaccination status prior to infection (Fully/Partially vs. Unvaccinated)[e] | **0.53 (0.07, 1.00)** | **0.57 (0.07, 1.07)** | 0.24 (-0.13, 0.61) | 0.24 (-0.14, 0.61) | 0.04 (-0.18, 0.26) | |
| Any medical care vs. None | **-0.94 (-1.57, -0.31)** | **-0.96 (-1.60, -0.31)** | -0.47 (-1.11, 0.16) | -0.43 (-1.07, 0.22) | 0.12 (-0.37, 0.60) | 0.09 (-0.40, 0.58) |
| Any symptom(s)[f] | -0.23 (-0.85, 0.40) | -0.22 (-0.85, 0.40) | -0.02 (-0.56, 0.52) | 0.04 (-0.52, 0.59) | 0.10 (-0.13, 0.33) | 0.09 (-0.14, 0.32) |
| ILI symptom(s)[f] | -0.38 (-0.86, 0.10) | -0.38 (-0.86, 0.11) | -0.34 (-0.71, 0.03) | -0.32 (-0.70, 0.06) | -0.05 (-0.27, 0.18) | -0.07 (-0.29, 0.16) |
| CLI symptom(s)[f] | -0.32 (-0.83, 0.19) | -0.33 (-0.84, 0.18) | -0.26 (-0.67, 0.16) | -0.23 (-0.66, 0.21) | 0.01 (-0.21, 0.23) | 0.00 (-0.22, 0.22) |
| Constitutional symptom(s)[f] | -0.42 (-0.95, 0.11) | -0.42 (-0.95, 0.11) | -0.30 (-0.72, 0.11) | -0.28 (-0.70, 0.13) | 0.02 (-0.20, 0.23) | 0.01 (-0.21, 0.23) |
| Moderate symptoms(s)[f] | -0.43 (-0.89, 0.04) | -0.43 (-0.90, 0.04) | **-0.38 (-0.76, -0.01)** | -0.36 (-0.75, 0.02) | -0.06 (-0.28, 0.17) | -0.07 (-0.29, 0.15) |
| Upper respiratory symptom(s)[f] | -0.10 (-0.62, 0.41) | -0.11 (-0.63, 0.41) | -0.10 (-0.58, 0.38) | -0.08 (-0.56, 0.40) | -0.00 (-0.22, 0.22) | -0.01 (-0.24, 0.21) |
| Lower respiratory symptom(s)[f] | -0.33 (-0.82, 0.16) | -0.35 (-0.84, 0.14) | -0.26 (-0.66, 0.15) | -0.23 (-0.65, 0.19) | 0.00 (-0.21, 0.22) | -0.00 (-0.22, 0.21) |
| Neurological symptom(s)[f] | -0.20 (-0.69, 0.30) | -0.19 (-0.70, 0.31) | -0.28 (-0.68, 0.13) | -0.24 (-0.68, 0.19) | 0.03 (-0.18, 0.25) | 0.02 (-0.20, 0.24) |
| Gastrointestinal symptom(s)[f] | **-0.74 (-1.29, -0.19)** | **-0.76 (-1.30, -0.21)** | -0.09 (-0.51, 0.33) | -0.05 (-0.48, 0.37) | -0.16 (-0.42, 0.11) | -0.16 (-0.42, 0.11) |

CI = Confidence Interval, AA = Associates of Arts, GED = General Educational Development, ILI = Influenza-like illness, CLI = COVID-like illness;
SARS-CoV-2 infection period = defined by the time of EQ-5D-3L survey after symptom onset/positive test
Adults = participants aged ≥16 years
[a] SARS-CoV-2 infection period observations not mutually exclusive
[b] Boldface indicates statistically significant results at the 0.05 level
[c] Significant findings may be underpowered due to small sample size
[d] Models restricted to participants aged ≥18 years
[e] Partially vaccinated: received 1 dose of 2 dose SARS-CoV-2 vaccine; Fully vaccinated: received 2 doses of 2 dose SARS-CoV-2 vaccine or 1 dose of 1 dose SARS-CoV-2 vaccine
[f] Referent group are participants who did not meet syndrome definition

Supplemental Table 5. Change in overall self-rated health status from enrollment among adults with SARS-CoV-2 infection, by infection period stratified by demographic/medical characteristics with >5 participants in each infection period

| | | | |
|---|---|---|---|
| **0–14-days (early) (n=61)[a]** | | | |
| | Vaccinated (Fully/Partially) n (column %) | Unvaccinated n (column %) | p-value[b] |
| No Change | 13 (37.1) | 9 (34.6) | **<0.01** |
| Improve | 10 (28.6) | 0 (0.0) | |
| Worsen | 9 (25.7) | 14 (53.8) | |
| Mixed Change | 3 (8.6) | 3 (11.5) | |
| | Reported seeking medical care during SARS-CoV-2 infection n (column %) | Reported seeking no medical care during SARS-CoV-2 infection n (column %) | p-value[b] |
| No Change | 1 (12.5) | 19 (37.3) | 0.22 |
| Improve | 0 (0.0) | 10 (19.6) | |
| Worsen | 7 (87.5) | 17 (33.3) | |
| Mixed Change | 0 (0.0) | 5 (9.8) | |
| | Reported gastrointestinal symptoms n (column %) | Reported no gastrointestinal symptoms n (column %) | p-value[b] |
| No Change | 2 (14.3) | 20 (42.6) | 0.06 |
| Improve | 1 (7.1) | 9 (19.1) | |
| Worsen | 9 (19.1) | 14 (29.8) | |
| Mixed Change | 2 (14.3) | 4 (8.5) | |
| **15–30 days (middle) (n=106)[a]** | | | |
| | Employed n (column %) | Unemployed n (column %) | p-value[b] |
| No Change | 44 (56.4) | 7 (29.2) | 0.07 |
| Improve | 12 (15.4) | 8 (33.3) | |
| Worsen | 20 (25.6) | 8 (33.3) | |
| Mixed Change | 2 (2.6) | 1 (4.2) | |
| **31–90 days (late) (n=298)[a]** | | | |
| | Reported underlying condition(s) n (column %) | Reported no underlying condition(s) n (column %) | p-value[b] |
| No Change | 80 (52.6) | 99 (67.8) | 0.06 |
| Improve | 34 (22.4) | 25 (17.1) | |
| Worsen | 30 (19.7) | 18 (12.3) | |
| Mixed Change | 8 (5.3) | 4 (2.7) | |

SARS-CoV-2 infection period = defined by the time of EQ-5D-3L survey after symptom onset/positive test
Adults = participants aged ≥16 years
[a] Among participants with EQ-5D-3L surveys at enrollment and SARS-CoV-2 infection period
[b] Boldface indicates significance at the 0.05 level

Supplemental Table 6. Descriptive statistics for health utilities among participants aged ≥8 years with SARS-CoV-2 infection, by infection period, stratified by age and reported presence of symptoms (restricted to 15-30-day and 31-90-day infection periods)

| SARS-CoV-2 infection period by self-report of symptoms[c] | No. | Mean (SD) | Median (IQR) | Bootstrapped 95% CI | Range |
|---|---|---|---|---|---|
| **Participants ≥16 years[a]** | | | | | |
| Reported Symptoms | | | | | |
| Overall | 341 | 0.90 (0.15) | 1.00 (0.83, 1.00) | 0.89,0.92 | 0.17-1.00 |
| 15-30 days (middle) | 101 | 0.87 (0.17) | 1.00 (0.81, 1.00) | 0.83,0.91 | 0.17-1.00 |
| 31-90 days (late) | 240 | 0.91 (0.13) | 1.00 (0.83, 1.00) | 0.90,0.93 | 0.17-1.00 |
| Reported No Symptoms | | | | | |
| Overall | 39 | 0.93 (0.12) | 1.00 (0.83, 1.00) | 0.89,0.96 | 0.45-1.00 |
| 15-30 days (middle) | 16 | 0.89 (0.15) | 0.93 (0.83, 1.00) | 0.81,0.95 | 0.45-1.00 |
| 31-90 days (late) | 23 | 0.95 (0.09) | 1.00 (0.92, 1.00) | 0.91,0.98 | 0.78-1.00 |
| **Participants 8-15 years[b]** | | | | | |
| SARS-CoV-2 infection period by self-report of symptoms[c] | No. | Mean (SD) | Median (IQR) | Bootstrapped 95% CI | Range |
| Reported Symptoms | | | | | |
| Overall | 78 | 0.94 (0.14) | 1.00 (0.89, 1.00) | 0.90,0.96 | 0.12-1.00 |
| 15-30 days (middle) | 21 | 0.88 (0.23) | 1.00 (0.89, 1.00) | 0.78,0.96 | 0.12-1.00 |
| 31-90 days (late) | 57 | 0.95 (0.08) | 1.00 (0.89, 1.00) | 0.93,0.97 | 0.71-1.00 |
| Reported No Symptoms | | | | | |
| Overall | 12 | 0.94 (0.10) | 1.00 (0.84, 1.00) | 0.87,0.99 | 0.71-1.00 |
| 15-30 days (middle) | 6 | 0.97 (0.06) | 1.00 (1.00, 1.00) | 0.92,1.00 | 0.84-1.00 |
| 31-90 days (late) | 6 | 0.90 (0.12) | 0.92 (0.83, 1.00) | 0.80,0.97 | 0.71-1.00 |

SD = standard deviation; IQR = interquartile range; Cl = confidence interval
SARS-CoV-2 infection period = defined by the time of EQ-5D-3L survey after symptom onset/positive test
[a] US value sets for adults (aged ≥16 years) (Shaw JW, Johnson JA, Coons SJ. US valuation of the EQ-5D health states: development and testing of the D1 valuation model. *Med Care*. Mar 2005;43(3):203-20. doi:10.1097/00005650-200503000-00003)
[b] Value sets used to calculate health utilities derived from Spanish children (aged 8-15 years) onset/positive test (Ramos-Goñi JM, Oppe M, Estévez-Carrillo A, et al. Accounting for Unobservable Preference Heterogeneity and Evaluating Alternative Anchoring Approaches to Estimate Country-Specific EQ-5D-Y Value Sets: A Case Study Using Spanish Preference Data. *Value in Health*. 2022;25(5):835-843. doi:10.1016/j.jval.2021.10.013)
[c] Self-reported at least one of the following ≤30 days post-onset/positive test: fever, cough, loss/change in taste/smell, sore throat, muscle/body aches, shortness of breath, diarrhea, fatigue, headache, nasal congestion, vomiting

Supplemental Table 7. Descriptive statistics for health utilities among participants aged ≥8 years with SARS-CoV-2 infection, by infection period, stratified by age, presence of symptoms, and vaccination status prior to infection (restricted to 15-30-day and 31-90-day infection periods)

| Participants ≥16 years[a] | | | | | |
|---|---|---|---|---|---|
| SARS-CoV-2 infection period by self-report of symptoms[c] and vaccination status[d] | No. | Mean (SD) | Median (IQR) | Bootstrapped 95% CI | Range |
| Reported Symptoms—Fully / Partially Vaccinated | | | | | |
| Overall | 223 | 0.91 (0.13) | 1.00 (0.83, 1.00) | 0.89,0.93 | 0.31-1.00 |
| 15-30 days (middle) | 58 | 0.90 (0.12) | 1.00 (0.83, 1.00) | 0.87,0.93 | 0.52-1.00 |
| 31-90 days (late) | 165 | 0.91 (0.13) | 1.00 (0.83, 1.00) | 0.89,0.93 | 0.31-1.00 |
| Reported No Symptoms—Fully / Partially Vaccinated | | | | | |
| Overall | 18 | 0.95 (0.09) | 1.00 (0.88, 1.00) | 0.90,0.98 | 0.78-1.00 |
| 15-30 days (middle) | 2 | 1.00 (0.00) | 1.00 (1.00, 1.00) | 1.00,1.00 | 1.00-1.00 |
| 31-90 days (late) | 16 | 0.94 (0.09) | 1.00 (0.84, 1.00) | 0.89,0.98 | 0.78-1.00 |
| Reported Symptoms—Unvaccinated | | | | | |
| Overall | 118 | 0.88 (0.18) | 1.00 (0.82, 1.00) | 0.85,0.91 | 0.17-1.00 |
| 15-30 days (middle) | 43 | 0.83 (0.21) | 0.84 (0.77, 1.00) | 0.77,0.89 | 0.17-1.00 |
| 31-90 days (late) | 75 | 0.91 (0.15) | 1.00 (0.84, 1.00) | 0.88,0.94 | 0.17-1.00 |
| Reported No Symptoms—Unvaccinated | | | | | |
| Overall | 21 | 0.91 (0.14) | 1.00 (0.83, 1.00) | 0.84,0.96 | 0.45-1.00 |
| 15-30 days (middle) | 14 | 0.88 (0.15) | 0.85 (0.83, 1.00) | 0.79,0.94 | 0.45-1.00 |
| 31-90 days (late) | 7 | 0.97 (0.08) | 1.00 (1.00, 1.00) | 0.91,1.00 | 0.80-1.00 |
| Participants 8-15 years[b] | | | | | |
| SARS-CoV-2 infection period by self-report of symptoms[c] and vaccination status[d] | No. | Mean (SD) | Median (IQR) | Bootstrapped 95% CI | Range |
| Reported Symptoms — Fully / Partially Vaccinated | | | | | |
| Overall | 33 | 0.95 (0.08) | 1.00 (0.89, 1.00) | 0.93,0.98 | 0.71-1.00 |
| 15-30 days (middle) | 5 | 1.00 (0.00) | 1.00 (1.00, 1.00) | 1.00, 1.00 | 1.00-1.00 |
| 31-90 days (late) | 28 | 0.94 (0.08) | 1.00 (0.89, 1.00) | 0.91,0.97 | 0.71-1.00 |
| Reported No Symptoms — Fully / Partially Vaccinated | | | | | |
| Overall | 6 | 0.97 (0.06) | 1.00 (1.00, 1.00) | 0.92, 1.00 | 0.84-1.00 |
| 15-30 days (middle) | 2 | 1.00 (0.00) | 1.00 (1.00, 1.00) | 1.00, 1.00 | 1.00-1.00 |
| 31-90 days (late) | 4 | 0.96 (0.08) | 1.00 (0.96, 1.00) | 0.88, 1.00 | 0.84-1.00 |
| Reported Symptoms — Unvaccinated | | | | | |
| Overall | 45 | 0.92 (0.17) | 1.00 (0.89, 1.00) | 0.87,0.96 | 0.12-1.00 |
| 15-30 days (middle) | 16 | 0.85 (0.25) | 1.00 (0.80, 1.00) | 0.72,0.95 | 0.12-1.00 |
| 31-90 days (late) | 29 | 0.96 (0.08) | 1.00 (1.00, 1.00) | 0.93,0.99 | 0.72-1.00 |
| Reported No Symptoms — Unvaccinated | | | | | |
| Overall | 6 | 0.90 (0.12) | 0.92 (0.83, 1.00) | 0.80,0.97 | 0.71-1.00 |
| 15-30 days (middle) | 4 | 0.96 (0.08) | 1.00 (0.96, 1.00) | 0.88, 1.00 | 0.84-1.00 |
| 31-90 days (late) | 2 | 0.77 (0.08) | 0.77 (0.74, 0.80) | 0.71,0.83 | 0.71-0.83 |

SD = standard deviation; IQR = interquartile range; Cl = confidence interval
SARS-CoV-2 infection period = defined by the time of EQ-5D-3L survey after symptom onset/positive test
[a] US value sets for adults (aged ≥16 years) (Shaw JW, Johnson JA, Coons SJ. US valuation of the EQ-5D health states: development and testing of the D1 valuation model. *Med Care*. Mar 2005;43(3):203-20. doi:10.1097/00005650-200503000-00003)
[b] Value sets used to calculate health utilities derived from Spanish children (aged 8-15 years) onset/positive test (Ramos-Goñi JM, Oppe M, Estévez-Carrillo A, et al. Accounting for Unobservable Preference Heterogeneity and Evaluating Alternative Anchoring Approaches to Estimate Country-Specific EQ-5D-Y Value Sets: A Case Study Using Spanish Preference Data. *Value in Health*. 2022;25(5):835-843. doi:10.1016/j.jval.2021.10.013)
[c] Self-reported at least one of the following ≤30 days post-onset/positive test: fever, cough, loss/change in taste/smell, sore throat, muscle/body aches, shortness of breath, diarrhea, fatigue, headache, nasal congestion, vomiting
[d] Partially vaccinated: received 1 dose of 2 dose SARS-CoV-2 vaccine; Fully vaccinated: received 2 doses of 2 dose SARS-CoV-2 vaccine or 1 dose of 1 dose SARS-CoV-2 vaccine

Supplemental Table 8. Descriptive statistics for health utilities among participants aged ≥8 years with SARS-CoV-2 infection, by infection period, stratified by age and illness severity (restricted to 15-30-day and 31-90-day infection periods)

| Participants ≥16 years[a] | | | | | |
|---|---|---|---|---|---|
| SARS-CoV-2 infection period by illness severity | No. | Mean (SD) | Median (IQR) | Bootstrapped 95% CI | Range |
| Asymptomatic[c] | | | | | |
| Overall | 39 | 0.93 (0.12) | 1.00 (0.83, 1.00) | 0.89,0.96 | 0.45-1.00 |
| 15-30 days (middle) | 16 | 0.89 (0.15) | 0.93 (0.83, 1.00) | 0.82,0.95 | 0.45,1.00 |
| 31-90 days (late) | 23 | 0.95 (0.09) | 1.00 (0.92, 1.00) | 0.91,0.98 | 0.78-1.00 |
| Mild[d] | | | | | |
| Overall | 312 | 0.91 (0.14) | 1.00 (0.83, 1.00) | 0.89,0.92 | 0.17-1.00 |
| 15-30 days (middle) | 90 | 0.89 (0.15) | 1.00 (0.81, 1.00) | 0.86,0.92 | 0.26-1.00 |
| 31-90 days (late) | 222 | 0.91 (0.14) | 1.00 (0.83, 1.00) | 0.89,0.93 | 0.17-1.00 |
| Severe[e] | | | | | |
| Overall | 29 | 0.86 (0.20) | 1.00 (0.82, 1.00) | 0.79,0.93 | 0.17-1.00 |
| 15-30 days (middle) | 11 | 0.76 (0.28) | 0.83 (0.63, 1.00) | 0.58,0.9 | 0.17-1.00 |
| 31-90 days (late) | 18 | 0.93 (0.11) | 1.00 (0.83, 1.00) | 0.88,0.98 | 0.72-1.00 |
| Hospitalized[f] | | | | | |
| Overall | 1 | 0.84 (NA) | 0.84 (0.84, 0.84) | - | 0.84-0.84 |
| 15-30 days (middle) | 0 | - | - | - | - |
| 31-90 days (late) | 1 | 0.84 (NA) | 0.84 (0.84, 0.84) | - | 0.84-0.84 |
| Participants 8-15 years[b] | | | | | |
| SARS-CoV-2 infection period by illness severity | No. | Mean (SD) | Median (IQR) | Bootstrapped 95% CI | Range |
| Asymptomatic[c] | | | | | |
| Overall | 12 | 0.94 (0.10) | 1.00 (0.84, 1.00) | 0.88,0.99 | 0.71-1.00 |
| 15-30 days (middle) | 6 | 0.97 (0.06) | 1.00 (1.00, 1.00) | 0.92,1.00 | 0.84-1.00 |
| 31-90 days (late) | 6 | 0.90 (0.12) | 0.92 (0.83, 1.00) | 0.8,0.97 | 0.71-1.00 |
| Mild[d] | | | | | |
| Overall | 75 | 0.94 (0.12) | 1.00 (0.89, 1.00) | 0.91,0.97 | 0.12-1.00 |
| 15-30 days (middle) | 19 | 0.92 (0.21) | 1.00 (0.94, 1.00) | 0.8,0.99 | 0.12-1.00 |
| 31-90 days (late) | 56 | 0.95 (0.08) | 1.00 (0.89, 1.00) | 0.93,0.97 | 0.71-1.00 |
| Severe[e] | | | | | |
| Overall | 3 | 0.70 (0.26) | 0.61 (0.56, 0.81) | 0.5,1.00 | 0.50-1.00 |
| 15-30 days (middle) | 2 | 0.56 (0.08) | 0.56 (0.53, 0.58) | 0.5,0.61 | 0.50-0.61 |
| 31-90 days (late) | 1 | 1.00 (NA) | 1.00 (1.00, 1.00) | - | 1.00-1.00 |
| Hospitalized[f] | | | | | |
| Overall | 0 | - | - | - | - |
| 15-30 days (middle) | 0 | - | - | - | - |
| 31-90 days (late) | 0 | - | - | - | - |

SD = standard deviation; IQR = interquartile range; Cl = confidence interval
SARS-CoV-2 infection period = defined by the time of EQ-5D-3L survey after symptom onset/positive test
[a] US value sets for adults (aged ≥16 years) (Shaw JW, Johnson JA, Coons SJ. US valuation of the EQ-5D health states: development and testing of the D1 valuation model. *Med Care*. Mar 2005;43(3):203-20. doi:10.1097/00005650-200503000-00003)
[b] Value sets used to calculate health utilities derived from Spanish children (aged 8-15 years) onset/positive test (Ramos-Goñi JM, Oppe M, Estévez-Carrillo A, et al. Accounting for Unobservable Preference Heterogeneity and Evaluating Alternative Anchoring Approaches to Estimate Country-Specific EQ-5D-Y Value Sets: A Case Study Using Spanish Preference Data. *Value in Health*. 2022;25(5):835-843. doi:10.1016/j.jval.2021.10.013)
[c] Self-reported none of the following symptoms ≤30 days post-onset/positive test: fever, cough, loss/change in taste/smell, sore throat, muscle/body aches, shortness of breath, diarrhea, fatigue, headache, nasal congestion, vomiting
[d] Self-reported at least one of the following symptoms ≤30 days post-onset/positive test: fever, cough, loss/change in taste/smell, sore throat, muscle/body aches, shortness of breath, diarrhea, fatigue, headache, nasal congestion, vomiting
[e] Self-reported non-hospitalized medical care ≤30 days post-onset/positive test
[f] Self-reported hospitalization ≤30 days post-onset/positive test

Supplemental Table 9. Descriptive statistics for health utilities among participants aged ≥8 years with SARS-CoV-2 infection, by infection period, stratified by age and cohort

| Participants ≥16 years[a] | | | | | |
|---|---|---|---|---|---|
| SARS-CoV-2 infection period by cohort | No. | Mean (SD) | Median (IQR) | Bootstrapped 95% CI | Range |
| PACC | | | | | |
| Overall | 470 | 0.91 (0.12) | 1.00 (0.83, 1.00) | 0.90,0.92 | 0.22-1.00 |
| 0-14 days (early) | 44 | 0.89 (0.15) | 1.00 (0.83, 1.00) | 0.85,0.93 | 0.22-1.00 |
| 15-30 days (middle) | 90 | 0.91 (0.13) | 1.00 (0.83, 1.00) | 0.88,0.93 | 0.45-1.00 |
| 31-90 days (late) | 336 | 0.91 (0.12) | 1.00 (0.83, 1.00) | 0.90,0.93 | 0.31-1.00 |
| C-HEaRT/SEARCh | | | | | |
| Overall | 68 | 0.80 (0.20) | 0.83 (0.77, 1.00) | 0.76,0.85 | 0.17-1.00 |
| 0-14 days (early) | 26 | 0.81 (0.16) | 0.81 (0.77, 0.96) | 0.75,0.87 | 0.44-1.00 |
| 15-30 days (middle) | 27 | 0.77 (0.23) | 0.81 (0.74, 1.00) | 0.68,0.85 | 0.17-1.00 |
| 31-90 days (late) | 15 | 0.86 (0.23) | 1.00 (0.83, 1.00) | 0.73,0.95 | 0.17-1.00 |
| Participants 8-15 years[b] | | | | | |
| SARS-CoV-2 infection period by cohort | No. | Mean (SD) | Median (IQR) | Bootstrapped 95% CI | Range |
| PACC | | | | | |
| Overall | 108 | 0.96 (0.07) | 1.00 (0.89, 1.00) | 0.94,0.97 | 0.71-1.00 |
| 0-14 days (early) | 16 | 0.95 (0.07) | 1.00 (0.89, 1.00) | 0.92,0.98 | 0.83-1.00 |
| 15-30 days (middle) | 21 | 0.98 (0.05) | 1.00 (1.00, 1.00) | 0.95,1.00 | 0.83-1.00 |
| 31-90 days (late) | 71 | 0.95 (0.08) | 1.00 (0.89, 1.00) | 0.93,0.97 | 0.71-1.00 |
| C-HEaRT/SEARCh | | | | | |
| Overall | 16 | 0.79 (0.25) | 0.81 (0.69, 1.00) | 0.66,0.90 | 0.12-1.00 |
| 0-14 days (early) | 8 | 0.88 (0.17) | 1.00 (0.72, 1.00) | 0.77,0.97 | 0.61-1.00 |
| 15-30 days (middle) | 6 | 0.64 (0.31) | 0.67 (0.53, 0.85) | 0.40,0.85 | 0.12-1.00 |
| 31-90 days (late) | 2 | 0.86 (0.20) | 0.86 (0.79, 0.93) | 0.71,1.00 | 0.71-1.00 |

SD = standard deviation; IQR = interquartile range; Cl = confidence interval

SARS-CoV-2 infection period = defined by the time of EQ-5D-3L survey after symptom onset/positive test

[a] US value sets for adults (aged ≥16 years) (Shaw JW, Johnson JA, Coons SJ. US valuation of the EQ-5D health states: development and testing of the D1 valuation model. *Med Care*. Mar 2005;43(3):203-20. doi:10.1097/00005650-200503000-00003)

[b] Value sets used to calculate health utilities derived from Spanish children (aged 8-15 years) onset/positive test (Ramos-Goñi JM, Oppe M, Estévez-Carrillo A, et al. Accounting for Unobservable Preference Heterogeneity and Evaluating Alternative Anchoring Approaches to Estimate Country-Specific EQ-5D-Y Value Sets: A Case Study Using Spanish Preference Data. *Value in Health*. 2022;25(5):835-843. doi:10.1016/j.jval.2021.10.013)

Supplemental Table 10. Descriptive statistics for health utilities among participants aged ≥8 years with SARS-CoV-2 infection, by infection period, stratified by age and time of survey completion

| Participants ≥16 years[a] | | | | | |
|---|---|---|---|---|---|
| SARS-CoV-2 infection period by time of EQ-5D-3L completion | No. | Mean (SD) | Median (IQR) | Bootstrapped 95% CI | Range |
| Enrollment[cd] | | | | | |
| Overall | 465 | 0.91 (0.10) | 1.00 (0.83, 1.00) | 0.90,0.92 | 0.26-1.00 |
| 0-14 days (early) | 61 | 0.91 (0.09) | 0.84 (0.83, 1.00) | 0.89,0.93 | 0.75-1.00 |
| 15-30 days (middle) | 106 | 0.91 (0.10) | 0.93 (0.83, 1.00) | 0.89,0.93 | 0.55-1.00 |
| 31-90 days (late) | 298 | 0.91 (0.11) | 1.00 (0.83, 1.00) | 0.90,0.93 | 0.26-1.00 |
| End of follow-up[c] | | | | | |
| Overall | 465 | 0.90 (0.12) | 1.00 (0.83, 1.00) | 0.89,0.91 | 0.31-1.00 |
| 0-14 days (early) | 62 | 0.91 (0.09) | 1.00 (0.83, 1.00) | 0.89,0.94 | 0.77-1.00 |
| 15-30 days (middle) | 99 | 0.90 (0.11) | 1.00 (0.83, 1.00) | 0.88,0.93 | 0.51-1.00 |
| 31-90 days (late) | 304 | 0.90 (0.12) | 1.00 (0.83, 1.00) | 0.89,0.91 | 0.31-1.00 |
| Participants 8-15 years[b] | | | | | |
| SARS-CoV-2 infection period by time of EQ-5D-3L completion | No. | Mean (SD) | Median (IQR) | Bootstrapped 95% CI | Range |
| Enrollment[cd] | | | | | |
| Overall | 114 | 0.96 (0.08) | 1.00 (0.89, 1.00) | 0.94,0.97 | 0.61-1.00 |
| 0-14 days (early) | 22 | 0.96 (0.06) | 1.00 (0.91, 1.00) | 0.94,0.99 | 0.83-1.00 |
| 15-30 days (middle) | 24 | 0.97 (0.07) | 1.00 (0.97, 1.00) | 0.94,0.99 | 0.83-1.00 |
| 31-90 days (late) | 68 | 0.95 (0.09) | 1.00 (0.89, 1.00) | 0.93,0.97 | 0.61-1.00 |
| End of follow-up[c] | | | | | |
| Overall | 103 | 0.95 (0.09) | 1.00 (1.00, 1.00) | 0.94,0.97 | 0.55-1.00 |
| 0-14 days (early) | 22 | 0.96 (0.08) | 1.00 (1.00, 1.00) | 0.93,0.99 | 0.71-1.00 |
| 15-30 days (middle) | 21 | 0.95 (0.10) | 1.00 (1.00, 1.00) | 0.90,0.99 | 0.67-1.00 |
| 31-90 days (late) | 60 | 0.95 (0.10) | 1.00 (1.00, 1.00) | 0.93,0.97 | 0.55-1.00 |

SD = standard deviation; IQR = interquartile range; Cl = confidence interval
SARS-CoV-2 infection period = defined by the time of EQ-5D-3L survey after symptom onset/positive test
Mean/median number of days between symptom onset/positive test and EQ-5D-3L completion: early: 8.34/8.50; middle: 23.40/23.00; late: 52.45/50.00
[a] US value sets for adults (aged ≥16 years) (Shaw JW, Johnson JA, Coons SJ. US valuation of the EQ-5D health states: development and testing of the D1 valuation model. *Med Care*. Mar 2005;43(3):203-20. doi:10.1097/00005650-200503000-00003)
[b] Value sets used to calculate health utilities derived from Spanish children (aged 8-15 years) onset/positive test (Ramos-Goñi JM, Oppe M, Estévez-Carrillo A, et al. Accounting for Unobservable Preference Heterogeneity and Evaluating Alternative Anchoring Approaches to Estimate Country-Specific EQ-5D-Y Value Sets: A Case Study Using Spanish Preference Data. *Value in Health*. 2022;25(5):835-843. doi:10.1016/j.jval.2021.10.013)
[c] Among participants with EQ-5D-3L completion at enrollment or end of-follow-up
[d] Values represent health utilities prior to symptom onset/positive-test

Supplemental Table 11. Descriptive statistics for visual analog scale scores among participants aged ≥8 years with SARS-CoV-2 infection, by infection period, stratified by age and time of completion

| Participants ≥16 years | | | | | |
|---|---|---|---|---|---|
| SARS-CoV-2 infection period by time of VAS completion | No. | Mean (SD) | Median (IQR) | Bootstrapped 95% CI | Range |
| Enrollment[ab] | | | | | |
| Overall | 465 | 83.55 (12.57) | 85.00 (80.00, 90.00) | 82.43,84.62 | 10-100 |
| 0-14 days (early) | 61 | 83.92 (9.62) | 85.00 (80.00, 90.00) | 81.43,86.26 | 60-100 |
| 15-30 days (middle) | 106 | 84.21 (11.29) | 85.00 (80.00, 90.00) | 81.72,86.18 | 31-100 |
| 31-90 days (late) | 298 | 83.24 (13.52) | 85.00 (80.00, 90.00) | 81.68,84.70 | 10-100 |
| End of follow-up[a] | | | | | |
| Overall | 465 | 82.64 (13.98) | 85.00 (76.00, 90.00) | 81.32,84.02 | 1-100 |
| 0-14 days (early) | 62 | 81.87 (15.46) | 85.00 (75.00, 90.00) | 77.93,85.42 | 11-100 |
| 15-30 days (middle) | 99 | 84.87 (10.73) | 87.00 (80.00, 90.50) | 82.75,86.84 | 50-100 |
| 31-90 days (late) | 304 | 82.07 (14.55) | 85.00 (75.00, 90.00) | 80.38,83.60 | 1-100 |
| Participants 8-15 years | | | | | |
| SARS-CoV-2 infection period by time of VAS completion | No. | Mean (SD) | Median (IQR) | Bootstrapped 95% CI | Range |
| Enrollment[ab] | | | | | |
| Overall | 114 | 94.99 (7.09) | 99.00 (90.25, 100.00) | 93.68,96.24 | 62-100 |
| 0-14 days (early) | 22 | 93.91 (6.81) | 95.00 (90.00, 100.00) | 90.91,96.64 | 75-100 |
| 15-30 days (middle) | 24 | 95.88 (5.81) | 99.00 (94.75, 100.00) | 93.37,98.04 | 80-100 |
| 31-90 days (late) | 68 | 95.03 (7.62) | 99.00 (94.75, 100.00) | 93.09,96.78 | 62-100 |
| End of follow-up[a] | | | | | |
| Overall | 103 | 89.79 (13.20) | 91.00 (87.50, 100.00) | 87.12,92.14 | 5-100 |
| 0-14 days (early) | 22 | 90.36 (10.77) | 90.00 (88.00, 100.00) | 85.64,94.45 | 60-100 |
| 15-30 days (middle) | 21 | 90.10 (10.02) | 91.00 (85.00, 98.00) | 85.95,93.95 | 62-100 |
| 31-90 days (late) | 60 | 89.47 (15.02) | 91.50 (87.50, 100.00) | 85.52,92.80 | 5-100 |

VAS = visual analog scale; SD = standard deviation; IQR = interquartile range; Cl = confidence interval
SARS-CoV-2 infection period = defined by the time of EQ-5D-3L survey after symptom onset/positive test
Mean/median number of days between symptom onset/positive test and VAS completion: early: 8.34/8.50; middle: 23.40/23.00; late: 52.45/50.00
[a] Among participants with VAS completion at enrollment or end of-follow-up
[b] Values represent health utilities prior to symptom onset/positive-test

Supplemental Figure 1. Correlation between health utilities and visual analog scale scores among participants with SARS-CoV-2, by infection period

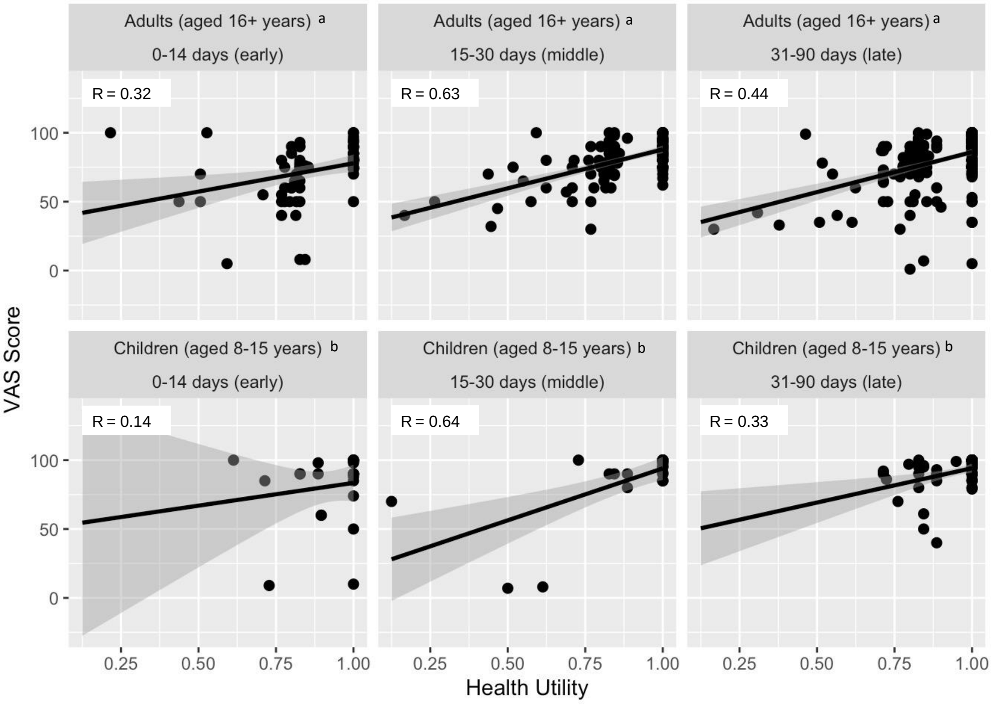


VAS = Visual Analog Scale
SARS-CoV-2 infection period = defined by the time of EQ-5D-3L survey after
[a] US value sets for adults (aged ≥16 years) (Shaw JW, Johnson JA, Coons SJ. US valuation of the EQ-5D health states: development and testing of the D1 valuation model. *Med Care*. Mar 2005;43(3):203-20.doi:10.1097/00005650-200503000-00003)
[b] Value sets used to calculate health utilities derived from Spanish children (aged 8-15 years) onset/positive test (Ramos-Goñi JM, Oppe M, Estévez-Carrillo A, et al. Accounting for Unobservable Preference Heterogeneity and Evaluating Alternative Anchoring Approaches to Estimate Country-Specific EQ-5D-Y Value Sets: A Case Study Using Spanish Preference Data. *Value in Health*. 2022;25(5):835-843.doi:10.1016/j.jval.2021.10.013)
